# Characteristics and Early Diagnosis of Motor Neuron Disease (MND) in 67 million individuals in England: a comparative study on phenotyping models derived by AI, Knowledge Graphs and the MND Association

**DOI:** 10.1101/2025.07.01.25330428

**Authors:** Yusuf Abdulle, Jinge Wu, Sanjay Budhdeo, Yunsoo Kim, Jiashu Shen, Emily Sun, Waqar Ali, Chengliang Dai, Phil Scordis, Arijit Patra, Ahmad Al Khleifat, Ammar Al-Chalabi, Alfredo Iacoangeli, Huanyu Zhang, Paul Taylor, Sarah Wild, Zina Ibrahim, Richard Dobson, Honghan Wu, the CVD-COVID-UK/COVID-IMPACT Consortium

## Abstract

**Background:** Motor neuron disease (MND) is a rapidly progressive and fatal neurodegenerative condition, making early diagnosis critical for optimizing patient outcomes and care planning. Despite the existence of decade-long clinical guidelines, early diagnosis of MND remains challenging due to the lack of population-level evidence on the effectiveness of what we know and, more importantly, what we do not know. This study aims to apply advanced computational methods on the whole English population linked health data to improve MND phenotype detection and diagnosis. Additionally, we assess the impact of COVID-19 on people with MND, examining mortality trends and vaccination effects.

**Methods:** The nationwide linked health records of 67 million individuals in England were used for identifying MND cohorts for two periods of 2014-2019 and 2020-2021, which were analysed to describe their characteristics and derive MND period prevalence. On this routinely collected health data, we implemented the MND Association’s red flag list (MNDA guideline) as well as three AI derived phenotyping models: knowledge graph, GPT-4 and machine learning on real-world data driven approaches. These phenotyping models were used in developing prediction models for (a) diagnosing MND; and (b) predicting MND 1, 3, and 5 years before coded diagnosis. Various computational methods were used in the implementation of prediction models including logistic regression, random forest, support vector machine as well as recurrent neural networks. The effectiveness was assessed using positive predictive value, sensitivity, F1 score, Area Under the Receiver Operating Characteristic Curve (AUROC) and specificity. The Kaplan-Meier method was used for conducting the survival analysis for COVID-19 related mortality of people with MND.

**Findings:** Of 67,270,015 individuals, from 1st January 2014 to 31st December 2019, we identified 12,240 people with coded MND diagnosis (6 year period prevalence of 18.20 cases per 100,000 people). For MND screening (task b), the MNDA guideline showed poor to moderate discrimination (AUROC: 0.62-0.63) while combining the guideline with AI derived ones (the ensemble) could improve it significantly (0.66-0.68). For the diagnosing task, the guideline had good discrimination (0.70), but lower overall performance (F1 score: 0.44) and the real-world data driven approach (hypothesis free) achieved much better results (AUROC: 0.78; F1: 0.55). As for COVID-19 mortality risk, compared to matched controls, MND patients had elevated risk (HR=2.97, 95% CI: 1.97-4.48) in wave 1, while fully vaccinated individuals in wave 2 demonstrated non-statistically significant higher risk (HR=1.27, 95% CI: 0.69-2.34).

**Interpretation:** This population scale study showed the MNDA guideline did not show very effective power in either screening or diagnosing MND probably due to the missing of predictive phenotypes available in routine care. The hypothesis-free phenotyping approach, applying AI on real-world datasets for deriving predictive phenotypes, demonstrated a great utility by identifying 13 novel phenotypes from 7 ICD-10 chapters that had significant effects in predicting MND.

**Funding:** British Heart Foundation Data Science Centre, led by Health Data Research UK. National Institute for Health and Care Research (NIHR) Dementia Biomedical Research Unit at South London and Maudsley NHS Foundation Trust and King’s College London.

## 1. Introduction

Motor Neuron Disease (MND) is a fatal neurodegenerative disease that has a projected age-adjusted incidence rate of 1.66 per 100,000 person-year.^1^ MNDs also comprise several subtypes, including amyotrophic lateral sclerosis (ALS), progressive bulbar palsy (PBP), primary lateral sclerosis (PLS) and progressive muscular atrophy (PMA). Characterised by progressive degeneration of upper and lower motor neurons, MND leads to debilitating impairments in bulbar, limb and diaphragmatic function. Affected individuals experience worsening dysphagia, dysarthria, respiratory issues, and motor dysfunction, ultimately resulting in significant morbidity and mortality.^2^ The incidence of MND peaks between the ages of 60 – 79 years..^3^ While some studies suggest a stabilisation in the incidence of the condition in the last couple of decades, others have reported an apparent increase in incidence, perhaps due to improved diagnosis and changes in the reporting standards. Given the global trend of population ageing, the burden of MND is anticipated to grow, reinforcing the urgency of early and accurate diagnosis to optimise patient care and access to therapeutic intervention.^2,3^.^5,6^

Accurate diagnosis of MND remains challenging, with multiple criteria proposed to improve diagnostic precision. The El Escorial Criteria, first introduced in 1994 and revised in 2000, were designed primarily for clinical trial recruitmnt and research standardisation.^7,8^ These were refined further in 2008 by the Awaji Criteria, which incorporated electrophysiological findings, including fasciculations, as key diagnostic markers of lower motor neuron involvement.^9,10^ More recently, they have been superseded by the Gold Coast criteria.^10^.^5^ Recognising the critical need for timely referrals, the Motor Neurone Disease Association (MNDA) collaborated with the Royal College of General Practitioners (RCGP) in 2014 to develop the MND Association Red Flag list—a tool aimed at facilitating earlier recognition and specialist referral from primary care.^11^ Early and accurate diagnosis is important for improving patient outcomes as this allows for timely access to multidisciplinary care and enrolment in clinical trials that use investigational therapies,^12^ but also provide people with MND and their families a stronger understanding of their anticipated disease course, medical decision-making and better planning.^13^

Clinical guidelines provide structured frameworks to enhance diagnostic consistency, yet their reliability is constrained by the quality of the available evidence, which may rely on expert opinion rather than robust data-driven insights.^14^ Moreover, there is limited transparency in guideline development methodologies, posing challenges for their applicability in diverse clinical settings. There is insufficient evidence on whether they work in real-world settings, the quantitative assessments of their performances in identifying people at different times, and, more importantly, whether there are better alternatives.

The availability of national linked health datasets (such as the whole English population linked health records) provides unprecedented opportunities in studying rare diseases. For example, a recent study on 331 rare diseases using such data is able to reveal novel knowledge at scale including new evidence of adjusted prevalence of 186 (56.2%) diseases they studied and 28 diseases with high risk of COVID-19 associated mortality. However, the detailed analysis of MND at the population level is missing, while the largest cohort study to date is 12,000.^15^

In parallel, biomedical research has increasingly adopted computational approaches—particularly machine learning (ML) and deep learning (DL)—to improve disease understanding and diagnosis.^16^ These techniques have been applied across electronic health records (EHRs),^17–26^ including studies on neurodegenerative diseases,^25–29^. There is an unparalleled opportunity to investigate how such advanced technologies can be applied to the abovementioned population EHRs for deriving novel insights for MND.

Another underutilzed resource for studying MND is the structured biomedical knowledge, also called Knowledge Graphs (KGs), which are usually curated from the scientific literature and publicly available databases.^32,33^ KGs represent rich, interconnected repositories linking diseases, symptoms, genes, and treatments, and have been successfully applied in clinical decision support, drug discovery, disease modelling,^34–36^ disease predictions,^37,38^ gene disease association analysis,^35,39–41^ as well as rare disease diagnosis. Similarly, recent breakthroughs of Generative Pre-trained Transformer (GPT) technologies are able to digest vast amounts, if not all, of human generated knowledge/information, aiming for artificial general intelligence, which holds a great potential for disease diagnosis for complexing diseases like MND.^42^

However, similar to the MND Red Flag List, such novel approaches have not been evaluated at scale to test their effectiveness in early diagnosis of MND, and indeed whether they offer an alternative or complementary mechanism to clinical guidelines. Utilising the whole English population linked health datasets^15^, this study aims to first identify and characterize MND subpopulation in England, and then bridge that gap by evaluating phenotypes derived from KGs, GPT-4^43^, the MNDA Red Flag List, and real-world data-driven approaches in the tasks of MND early diagnosis at varying time points—1, 3, and 5 years before coded diagnosis. We hypothesise that ensembled lists, which combine computationally derived phenotypes with the MNDA Red Flag List, will outperform individual methodologies in MND prediction. By integrating machine learning, knowledge graphs, and clinical guidelines, this research seeks to establish a more comprehensive and validated MND diagnostic framework. If successful, this approach could enhance early detection, facilitate targeted interventions, and ultimately improve patient outcomes. We will also analyze the risk of MND patients in COVID-19 associated mortality to understand their vulnerabilities and provide evidence for informing public health policies.

## 2. Methods

### 2.1 Study population and period

We carried out a retrospective observational cohort study across England, using nine connected datasets available within NHS England’s Secure Data Environment (SDE), accessed via the British Heart Foundation (BHF) Data Science Centre’s CVD-COVID-UK/COVID-IMPACT Consortium. These datasets included: primary care records from the General Practice Extraction Service for Pandemic Planning and Research (GDPPR);^44^ COVID-19 testing results from Public Health England’s Second Generation Surveillance System; hospital data from the Secondary Uses Service, including inpatient care (HES-APC), adult critical care, and outpatient services (HES-OP); hospitalisation records from the COVID-19 Hospitalisations in England Surveillance System; information on COVID-19 vaccination; and death registrations from the Office for National Statistics. The datasets contained only structured information—unstructured clinical notes and imaging were not included. As previously documented by Wood et al,^45^ this linked data resource reliably mirrors the demographic characteristics of the English population—such as age, sex at birth, and ethnicity— when compared with official government statistics.

This study utilised electronic health records (EHRs) for two separate cohorts. Firstly, we aimed to identify and characterise phenotypic profiles associated with an early diagnosis of MND using a historical cohort. Secondly, we established a contemporary cohort to explore factors associated with COVID-19 survival in patients with MND.

The study integrates healthcare data sources (GDPPR, HES-APC, and HES-OP) with a knowledge graph (PrimeKG) and machine learning approaches to identify and rank MND-associated phenotypes. Ranking algorithms—including graph machine learning methods, graph ranking algorithms, and GPT-4—are compared against clinical guidelines and real-world data. Predictive models (LSTM, logistic regression, SVM, and random forest) assess MND risk across 1-, 3-, and 5-year time windows prior to diagnosis. Evaluation metrics include F1 score, precision, recall, and AUROC. A separate COVID-19 analysis was conducted using a subset of the MND cohort from 2020–2022 to estimate survival during the pandemic, evaluated using Kaplan-Meier methods.

#### 2.1.1 Base Cohort

Patient data were retrospectively collected for the period 1 January 2014 to 31 December 2019. This cohort included individuals registered within the English National Health Service (NHS) who were alive after 1 January 2015, and subsequently received a diagnosis of MND between 1 January 2015 and 31 December 2019. MND cases were identified using specific diagnostic codes: the Systematised Nomenclature of Medicine – Clinical Terms (SNOMED-CT) code ‘86044005’ from primary care records and the International Classification of Diseases, Tenth Revision (ICD-10) code ‘G12.2’ from secondary care records. Each patient with MND was matched with three individuals without an MND diagnosis.

#### 2.1.2 Case and control identification

We included patients alive on 1 January 2015 with a first diagnosis of MND—SNOMED-CT “86044005” in GDPPR or ICD-10 “G12.2” in HES APC—between 1 January 2015 and 31 December 2019, and no death record before 1 January 2014. Each MND case was matched 1:3 to controls by age group (under 18, 18–29, 30–49, 50–69, ≥70 years), sex, and ethnicity (White, Asian/Asian British, Black/Black British, Mixed, Other) using stratified random sampling within each stratum.

#### 2.1.3 Temporal feature construction

We defined each case’s index date as their earliest MND code and generated feature matrices including all clinical codes occurring >1, >3, or >5 years before diagnosis. For controls, all clinical codes occurring before January 1, 2019 were extracted chronologically, with controls having any MND-related codes excluded. Where the pool of eligible controls exceeded demand, random sampling preserved the 1:3 ratio.

#### 2.1.4 COVID-19 Cohort

We performed a retrospective cohort analysis comparing COVID-19-related mortality in people with MND to matched controls from the general population. The study period spanned from September 2020 to November 2021, encompassing Wave 2 and the first half of Wave 3 of the UK pandemic (driven by the original strain/Alpha variant and Delta variant, respectively). This period was selected to reflect established vaccination rollout and high testing capacity.

Individuals were included if their first COVID-19 event occurred within the analysis period. For each MND patient, we identified three controls using exact matching on age group, sex, ethnicity, and vaccination status. This approach enabled precise comparison while accounting for key demographic confounders.

We identified COVID-19 events using five previously defined phenotypes: 1) positive SARS-CoV-2 tests, 2) COVID-19 diagnosis recorded in primary care, 3) hospital admissions with COVID-19 diagnosis, 4) ventilatory support related to COVID-19, and 5) COVID-19 mortality. Mortality included: a) confirmed or suspected COVID-19 diagnosis listed on the death certificate, b) death within 28 days of the first recorded COVID-19 event without COVID-19 on the death certificate, or c) COVID-19 hospital admission with discharge status indicating death. These phenotypes were identified using clinical codes from multiple linked datasets (SGSS, GDPPR, SUS, HES-APC, HES-CC, CHESS, ONS).

Vaccination status was determined from the COVID-19 vaccination dataset, with patients classified as fully vaccinated 14 days after their second dose.

### 2.2 Phenotyping for MND Prediction and Diagnosis

We used a diverse set of methods for deriving phenotyping models for two tasks of ascertaining MND diagnosis: prediction (predicting MND 1, 3 or 5 years before coded diagnosis) and diagnosis (identifying MND patients from the study cohort using all data available at coded diagnosis).

#### 2.2.1 Phenotypes from Clinical Guideline

As for a reference model, we used the Red Flag Diagnosis tool, developed by the MND Association in tandem with the RCGP. This tool, designed to speed up referrals to neurology and improve diagnostic accuracy, is tailored to identify early MND indicators in primary care. Using this tool, we obtained a curated list of phenotypes commonly associated with early stage MND based on clinical expertise. Appendix Table 1 shows the 20 phenotypes listed in the Red Flag Diagnosis tool ^(11,46)^.

**Table 1:**
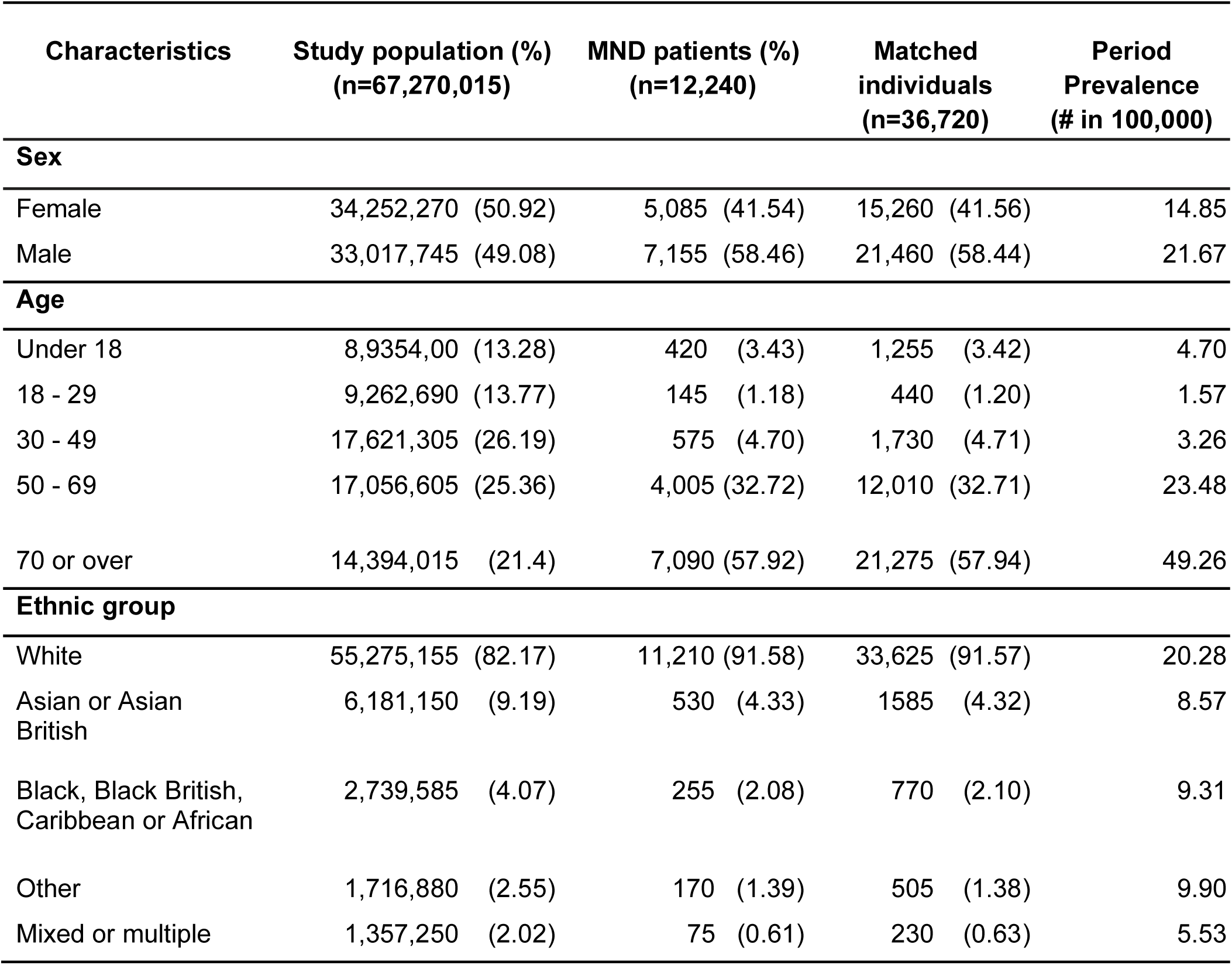
Demographic Characteristics of the UK Population and Individuals Diagnosed with Motor Neurone Disease (MND). This table presents the demographic distribution of the general UK population compared to individuals diagnosed with MND. Key characteristics include age, gender, ethnicity, alongside average survival years from diagnosis as well as average time from diagnosis to first symptom. The comparison highlights differences between the two groups, providing insights into potential risk factors and disparities in MND prevalence.

#### 2.2.2 Phenotypes from Knowledge Graph

To derive MND phenotypes from structured biomedical knowledge, we employed PrimeKG, a comprehensive knowledge graph (KG) designed for precision medicine, containing information from 20 biomedical resources and encompassing 17,080 diseases and over 4 million relationships.^33^ To focus our analysis on MND-relevant phenotypes, we extracted a subgraph extending two hops from the central MND node. Self-loops and isolated nodes were removed to enhance computational efficiency. The optimal subgraph size was determined by comparing MND subgraphs with Hepatitis subgraphs (as a control disease group with distinct pathophysiology and clinical presentation) at varying hop distances (K1 to K4, representing one to four hops). A few graph algorithms were used to derive the centralities (a quantification of importance) of nodes for each subgraph level: PageRank and Hyperlink-Induced Topic Search (HITS), ^47,48^ Node2Vec,^49^ Graph Convolutional Networks (GCNs),^50^ and Graph Transformers. Detailed descriptions of these algorithms are provided in the supplementary method section in the appendix.

#### 2.2.3 Phenotypes derived by Generative AI

OpenAI’s GPT-4 (March 14th, 2023 version) was used as a representative pretrained GPT model to populate a phenotyping model. It was prompted to list the top 20 phenotypes most commonly observed in patients with MND. This approach leveraged the model’s ability to synthesise and prioritise domain-relevant concepts without requiring access to structured datasets.

#### 2.2.4 Phenotyping models derived from Real World EHR

With the base MND cohort, we applied computational methods to derive phenotypes for MND. Using the Human Phenotype Ontology (HPO), we started with selecting all terms under its “Phenotypic abnormality” branch (HP:0000118) as the set of all potential abnormal phenotypes, yielding 16,798 distinct terms. Among these, 3,438 HPO terms were mapped, as defined in HPO, to 4,369 unique SNOMED-CT codes and 29 unique ICD-10 codes. We then implemented feature selection algorithms to obtain the most informative phenotypes for the prediction task on our MND cohort. The following algorithms were used for choosing the most predictive phenotypes: LASSO regularization with cross-validation, logistic regression with L1 penalty, correlation analysis, Chi-squared testing, and mutual information criteria. The top 20 phenotypes were selected to ensure direct comparability with the clinically curated Red Flag List.

### 2.3 The evaluation of phenotyping models in MND diagnosis and prediction

Two types of tasks were used to evaluate phenotyping models for ascertaining MND: prediction task and diagnosis task. The prediction task is to predict which individuals will develop MND in the future (1, 3 and 5 years before coded diagnosis). The diagnosis task is essentially to classify who has MND in the base cohort (i.e., using full history at the point of coded diagnosis). Five-fold cross validation was used for evaluation. Performance metrics included accuracy, Area Under the Receiver Operating Characteristic Curve (ROC-AUC), Positive Predictive Value (PPV), Sensitivity, Specificity, F1-score, as well as the confusion matrix. Feature importance (or attention weights) were assessed to identify most predictive phenotypes for MND. Four computational algorithms were implemented, ranging from statistical methods to deep learning ones, and the results of the best model (Random Forest) were used in the analysis. Details of all algorithms and technical specifications are available in the appendix. Phenotype importance for MND (early) diagnosis was assessed using odds ratio for logistic regression models and the SHAP (SHapley Additive exPlanations) approach for machine learning models(^51^). SHAP is a model agnostic approach to quantifying feature contributions to predictions.

### 2.4 Phenotyping Models’ Correlations and Statistical Analysis

To identify the added values of various computational methods, especially those leading to good MND ascertaining accuracies, we conducted comparative analyses of phenotyping models against the reference model (the MNDA Red Flag List) as well as between themselves. Specifically, Kendall’s Tau correlation was used to evaluate ordinal agreement between ranked phenotype lists, with values ranging from complete disagreement (−1) to perfect agreement (+1) and statistical significance threshold set at p<0.05. Additionally, the degree of phenotype set overlap was assessed by Jaccard similarity index, ranging from no overlap (0) to complete concordance (1), with p<0.05 indicating significant similarity.

### 2.5 COVID-19 Mortality Risk Analysis

Survival functions were estimated using the Kaplan-Meier method. Differences in survival distributions between MND patients and matched controls were assessed using the log-rank test. Hazard ratios (HRs) with 95% confidence intervals were estimated using univariable Cox proportional hazards models, with time of event as the dependent variable and MND status as the independent variable. The proportional hazards assumption was tested using Schoenfeld residuals.

To comprehensively assess the impact of vaccination, we stratified our analysis into four distinct cohorts: 1) overall cohort, 2) Wave 1 cohort, 3) Wave 2 non-vaccinated cohort, and 4) Wave 2 fully vaccinated cohort. This stratification allowed us to evaluate how vaccination status modified COVID-19 outcomes in MND patients.

## 3. Results

### 3.1 Population Statistics and MND Prevalence

Our study population included 67,270,015 individuals, registered with a GP or admitted into secondary care in England. From this population, using the base cohort definition, we identified 12,240 individuals with a coded Motor Neurone Disease (6 year period prevalence of 18.20 cases per 100,000 people, see Table 1 for details). Compared to the general population, this MND sub-population has higher proportions of males (58.46% in people with MND vs 49.08% in the study population), of people over the age of 50 (57.92% vs 25.36%) and over 70 (57.92% vs 21.4%). As for ethnicity, white individuals constitute a great proportion (91.58% vs 82.17% in the general population).

### 3.2 Phenotyping Models’ Performances in Diagnosing and Predicting MND

For the MND diagnosis task (using all phenotypes available prior to coded diagnosis), as seen in Table 2, the clinical guideline (MNDA) achieved an AUROC of 0.703 and an F1 score of 0.435. It was the second worst by AUROC with only GPT4 being worse. Its F1 score was the 3rd out of five models. Overall, the ensemble approach, combining MNDA Red Flag list, Knowledge Graph (KG) and the real world data driven approach (RWD), consistently performed the best on both AUROC (0.725) and F1 score (0.557). Compared to the MNDA Red Flag list, the ensemble approach significantly increased the F1 score (+0.122, 28%). For individual models, the RWD performed the second best on the two metrics. As for PPV, both MNDA and GPT4 models showed good performances, however, both of which had significant low sensitivity scores, missing high proportions (65.4% and 71.8%) of MND patients.

**Table 2:**
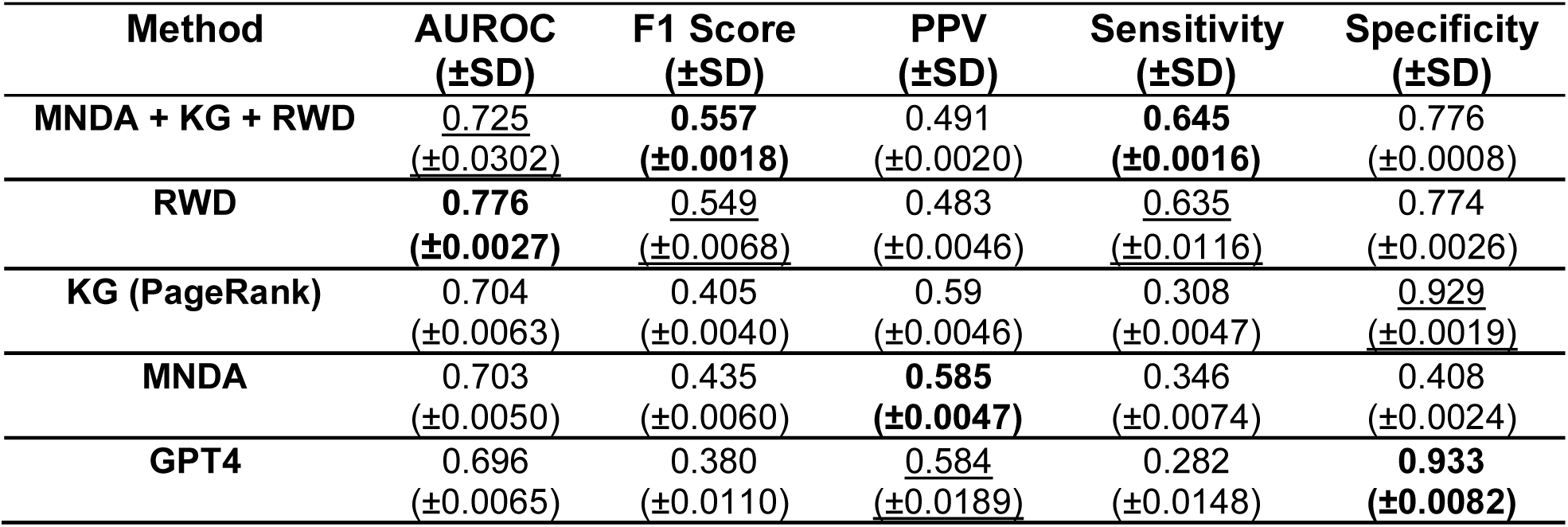
Performances of different phenotyping models on diagnosing MND. The method column indicates the derivation methods: MNDA → MNDA Red Flag List; RWD → logistic regression based feature selection on real world data; KG (PageRank) → PageRank algorithm derived from PrimeKG; GPT4 → generated by GPT-4; MNDA + KG + RWD → the ensemble method using MNDA, KG and RWD. The models are evaluated using AUROC (the area under the receiver operating characteristic curve), F1 score, PPV (Positive Predictive Value), Sensitivity, and Accuracy. Bold text indicates best performance while underlined ones are the second best.

| Method | AUROC<br>(±SD) | F1 Score<br>(±SD) | PPV<br>(±SD) | Sensitivity<br>(±SD) | Specificity<br>(±SD) |
| --- | --- | --- | --- | --- | --- |
| **MNDA + KG + RWD** | <u>0.725</u><br>(±0.0302) | **0.557**<br>(±0.0018) | 0.491<br>(±0.0020) | **0.645**<br>(±0.0016) | 0.776<br>(±0.0008) |
| **RWD** | **0.776**<br>(±0.0027) | <u>0.549</u><br>(±0.0068) | 0.483<br>(±0.0046) | <u>0.635</u><br>(±0.0116) | 0.774<br>(±0.0026) |
| **KG (PageRank)** | 0.704<br>(±0.0063) | 0.405<br>(±0.0040) | 0.59<br>(±0.0046) | 0.308<br>(±0.0047) | <u>0.929</u><br>(±0.0019) |
| **MNDA** | 0.703<br>(±0.0050) | 0.435<br>(±0.0060) | **0.585**<br>(±0.0047) | 0.346<br>(±0.0074) | 0.408<br>(±0.0024) |
| **GPT4** | 0.696<br>(±0.0065) | 0.380<br>(±0.0110) | <u>0.584</u><br>(±0.0189) | 0.282<br>(±0.0148) | **0.933**<br>(±0.0082) |

For MND prediction tasks (early diagnosis), as detailed in appendix tables S4-S6 for 1 year, 3 year and 5 year predictions respectively, the ensemble approach consistently performed the best on AUROCs (0.66 for 1 year, 0.67 for 3 year and 0.68 for 5 year) as well as on F1 scores (0.48, 0.49 and 0.50). The MNDA guideline did not perform as well on AUROCs (0.62, 0.62 and 0.63) and F1 scores (0.43, 0.43 and 0.44). The real world data approach again performed the second best overall and the best among the individual models with AUROCs of (0.65, 0.65 and 0.64) and F1 scores of (0.47, 0.48 and 0.47). For all early diagnosis tasks, the ensemble approach significantly outperformed the clinical guideline, with the biggest improvement on the 5 year early diagnosis task: 8% increase on AUROC and 14% on F1 score.

### 3.3 Important Phenotypes for Predicting and Diagnosing MND

Figure 2 gives the important phenotypes derived from the best performing models for both MND diagnosis and 3-year prediction. The sub figure Figure 2.a compares and contrasts the phenotypes at ICD-10 chapter level between individual models (knowledge graph, the MNDA guideline and real world data) in the ensemble approach. For the MND diagnosis task (the upper part of Figure 2a), all three individual models used chapter XVIII (Symptoms, signs and abnormal clinical and laboratory findings, not elsewhere classified) and VI (Diseases of the nervous system) with the former being ranked very high in all models. The MNDA guideline and knowledge graph approaches had more commonalities (chapters XVIII, XIII, VI and XXI). The RWD model used five extra chapters with IV (Endocrine, nutritional and metabolic diseases) and IX (Diseases of the circulatory system) being the second and third most important in its ranked list, respectively. For the 3 year MND prediction task (the lower part of Figure 2a), similarly, the knowledge graph model used all four chapters of the MNDA guideline with very similar rankings. The RWD model identified chapters XVIII, IX, V, IV and VI as the most predictive for the MND early diagnosis (3 years).

**Figure 1:**
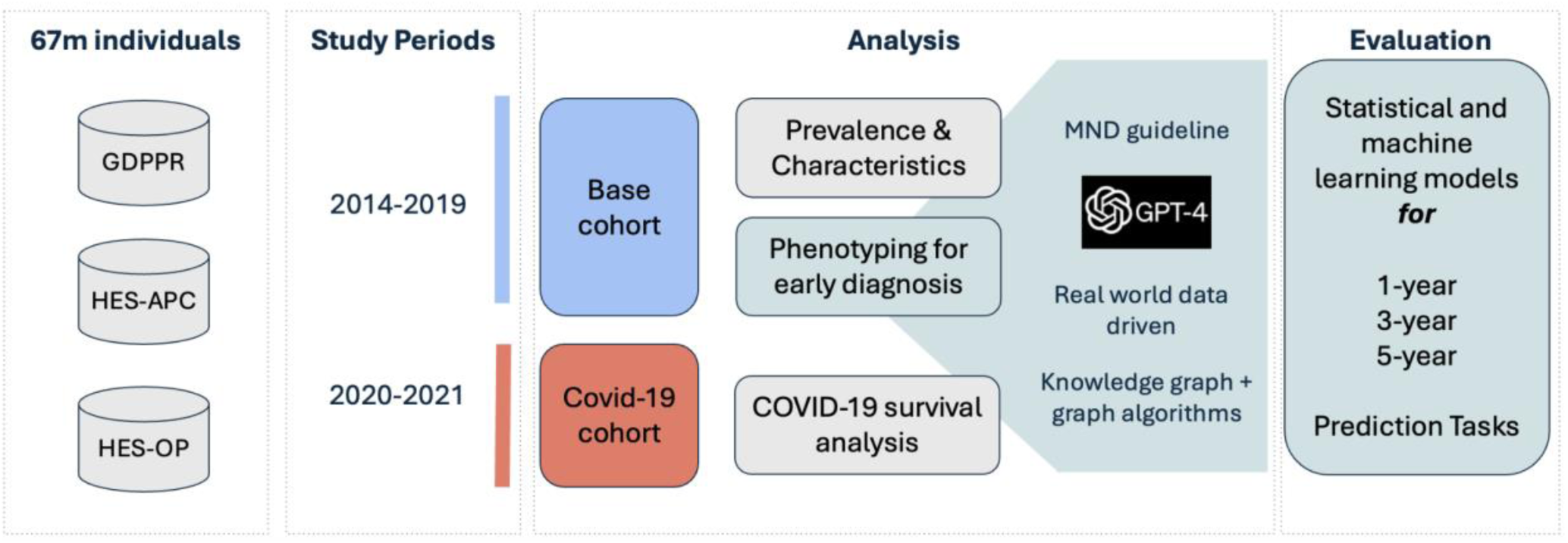
Overview of the methodological framework for MND phenotype ranking, prediction, and survival analysis.

**Figure 2:**
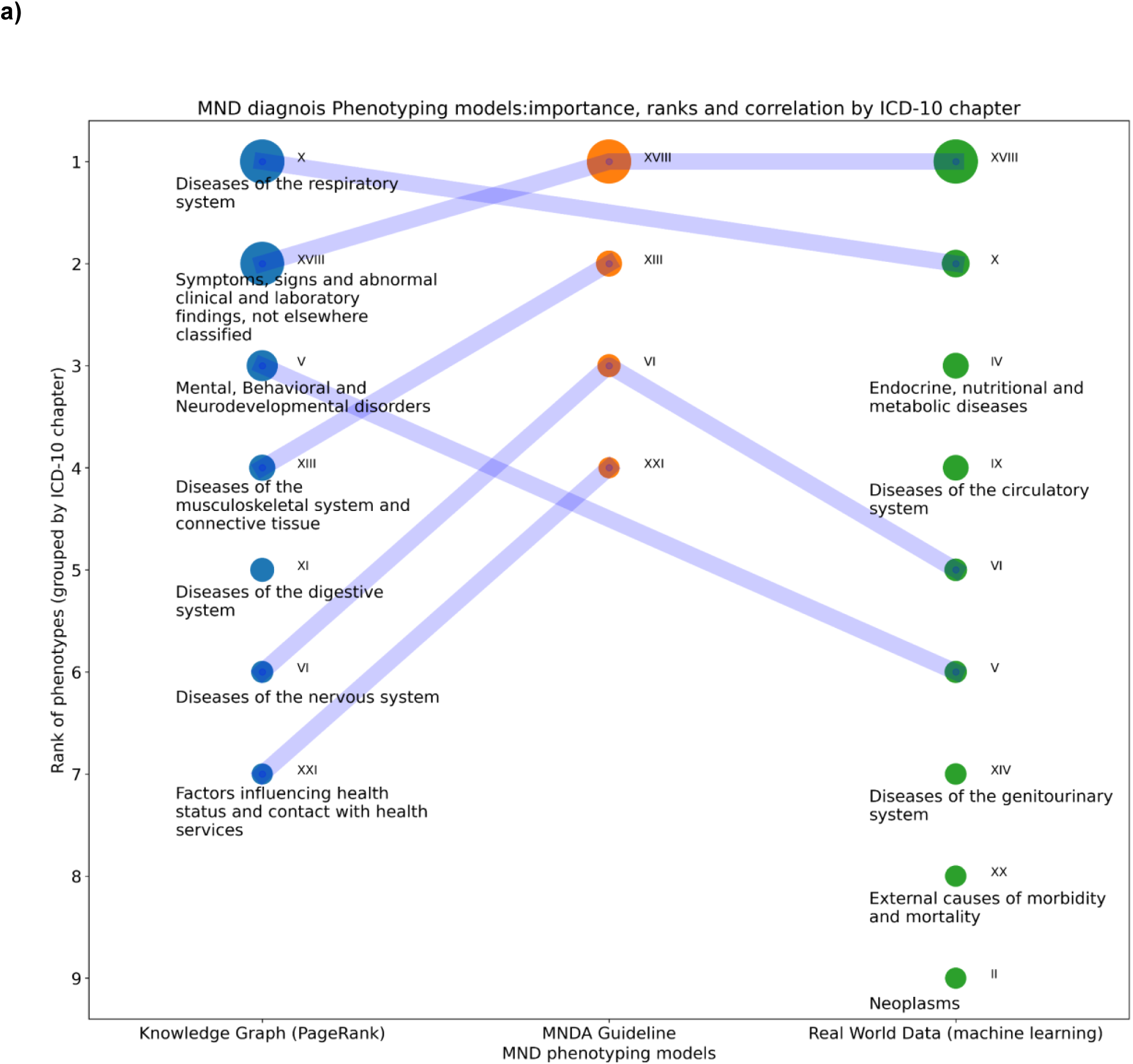
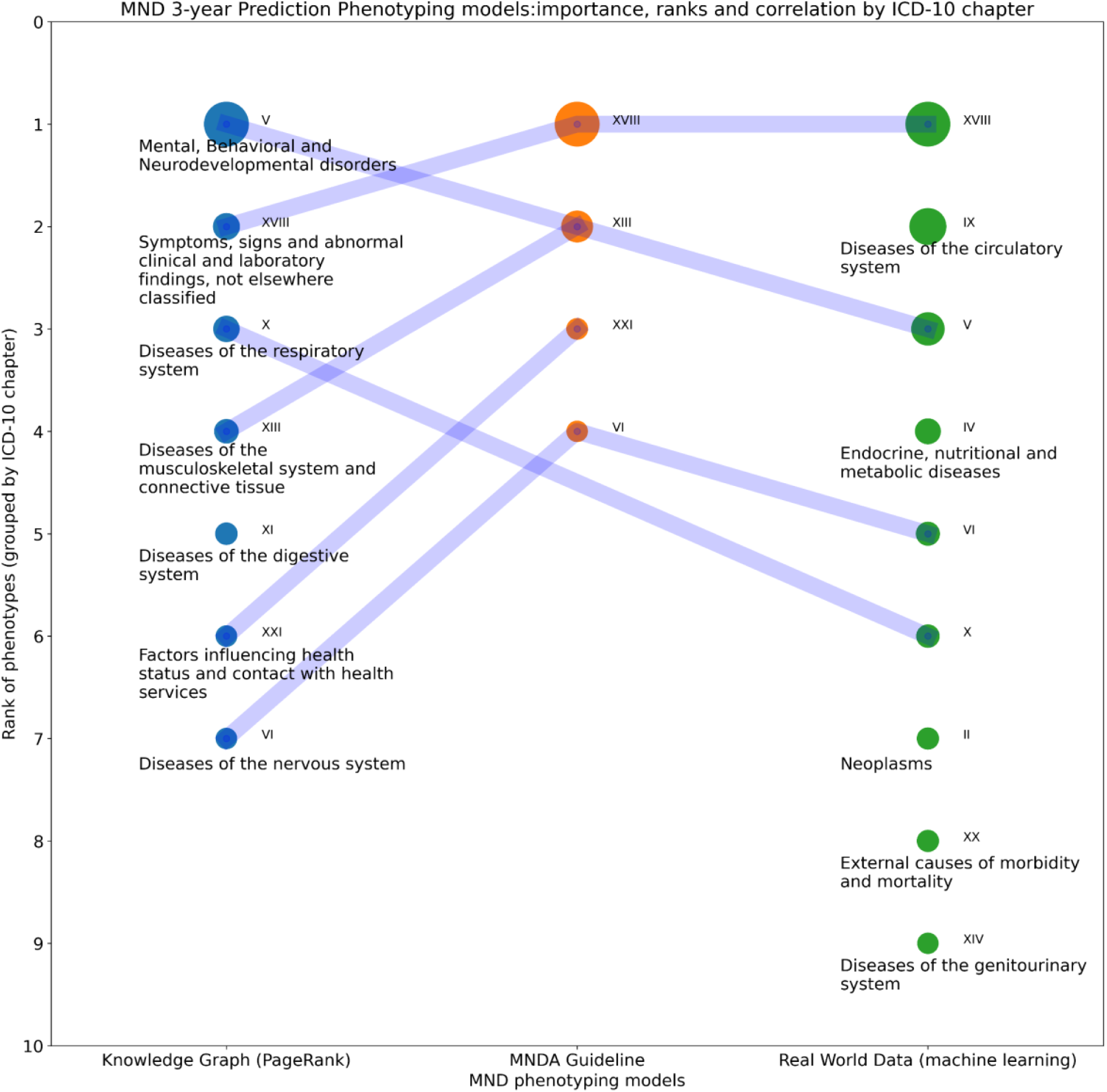
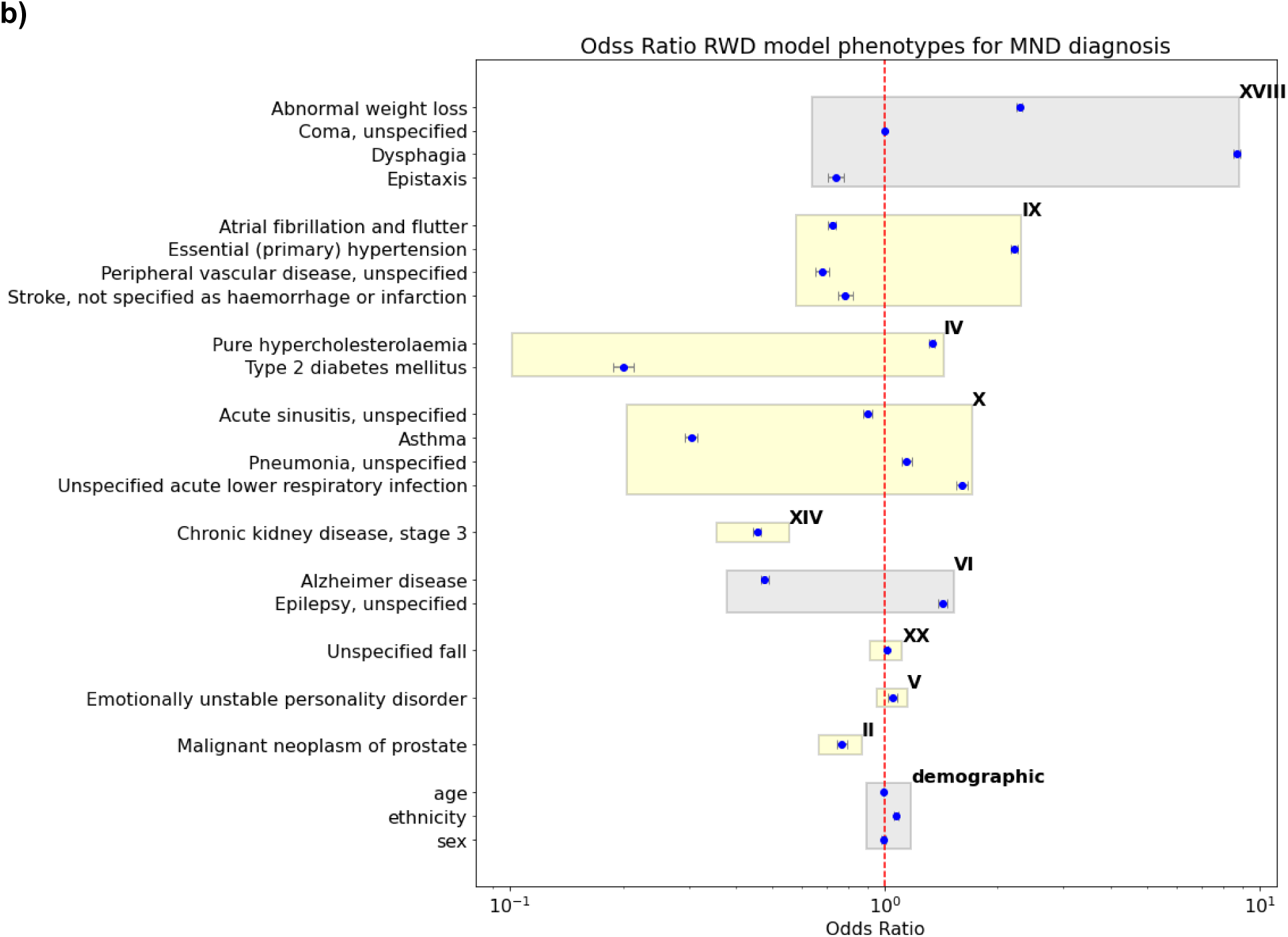
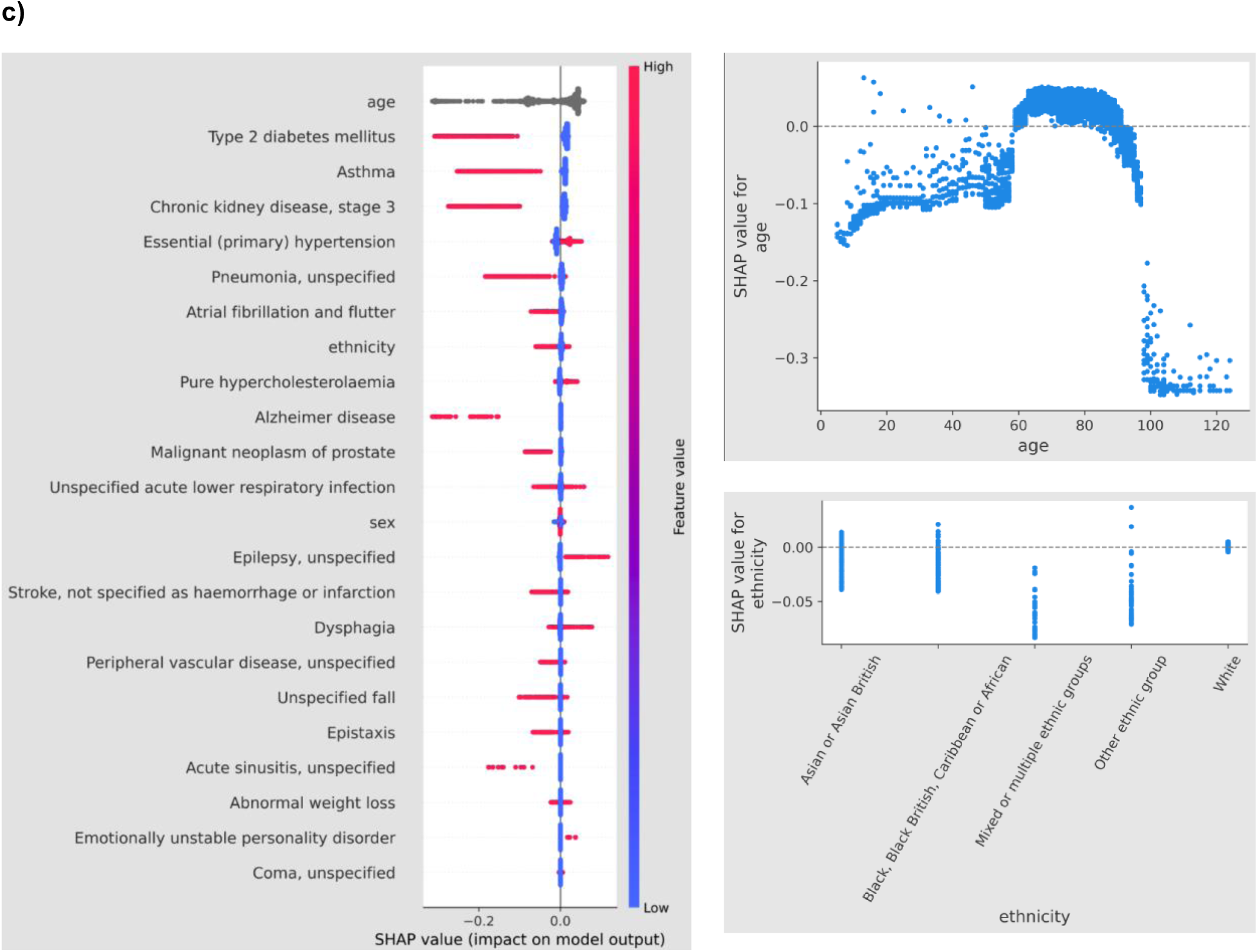
MND phenotyping models for diagnosis and prediction. **a)** Representative phenotyping models in the ensemble approach for MND diagnosis (upper) and 3-year MND prediction (lower). Phenotypes are grouped by ICD-10 chapters to give a high-level view. X-axis shows the different models. Y-axis is the rank of ICD-10 chapter as ordered by their importance (1 being most important). The size of the dots indicates the accumulated quantity of importance returned by the Random Forest model for diagnosing MND. The link between dots shows the commonality of ICD-10 chapters between models. **b)** The odds ratios of the (logistic regression based, AUROC=0.742) RWD model for MND diagnosis. The X-axis is the odds ratio in log-scale. The Y-axis is the list of phenotypes, which are grouped by ICD-10 chapters. The ICD-10 chapters are ordered by their accumulative importances (by the Random Forest model). The yellow background highlights those chapters that are not included in the MNDA guideline. For ethnicity, white is 1 with others being 0. **c)** The phenotype importance of the 3-year prediction RWD model (Random Forest, AUROC=0.766). The importance was assessed by SHAP values. The higher its absolute SHAP value, the more important (impactful) the phenotype is. Each dot represents an individual.

To investigate the importance of individual phenotypes, the RWD model (via feature selection on the real-world MND cohorts) was used for the analysis because it was the best individual phenotyping model for all diagnosis and prediction tasks. Figure 2b is the forest plot of odds ratios of phenotypes for diagnosing MND derived from a logistic regression model with AUROC=0.742. Phenotypes are grouped into boxes based on their ICD-10 chapters. Those with yellow background highlight phenotype categories that are not included in the MNDA guideline. Having Dysphagia (difficulty swallowing) and abnormal weight loss are associated with the highest risks of MND, followed by Essential hypertension and Unspecified acute lower respiratory infection. Type 2 diabetes mellitus and asthma are the most protective conditions, followed by Chronic kidney disease (stage 3) and Alzheimer disease. All phenotypes except hypertension in chapter IX (Diseases of the circulatory system) demonstrate protective effects.

For the 3-year early diagnosis task, Figure 2c shows the phenotype importance using SHAP method for all phenotypes as well as three demographic features of age, sex and ethnicity. The list is ordered by the impact of features. Age is the most predictive, which, however, has a complex effect pattern as shown in the dependency plot on the top right. For the rage from 60 to 90, age is shown to be associated with higher risks, while other ranges show protective effects. High risk phenotypes include Essential hypertension, Pure hypercholesterolaemia, Dysphagia and Epilepsy, unspecified. Type 2 diabetes mellitus and Asthma show most protective effects. For ethnicity, the bottom right subfigure shows that being in a minority group has a protective effect. Detailed analysis of other phenotype models are available in the supplementary results section in the appendix.

### 3.4 COVID-19 Mortality in People with MND

In the COVID-19 mortality analysis (Figure 3), MND patients demonstrated significantly higher COVID-19-related mortality compared to matched controls (log-rank p<0.0001), with a hazard ratio of 2.40 (95% CI: 1.88-3.06).

**Figure 3.**
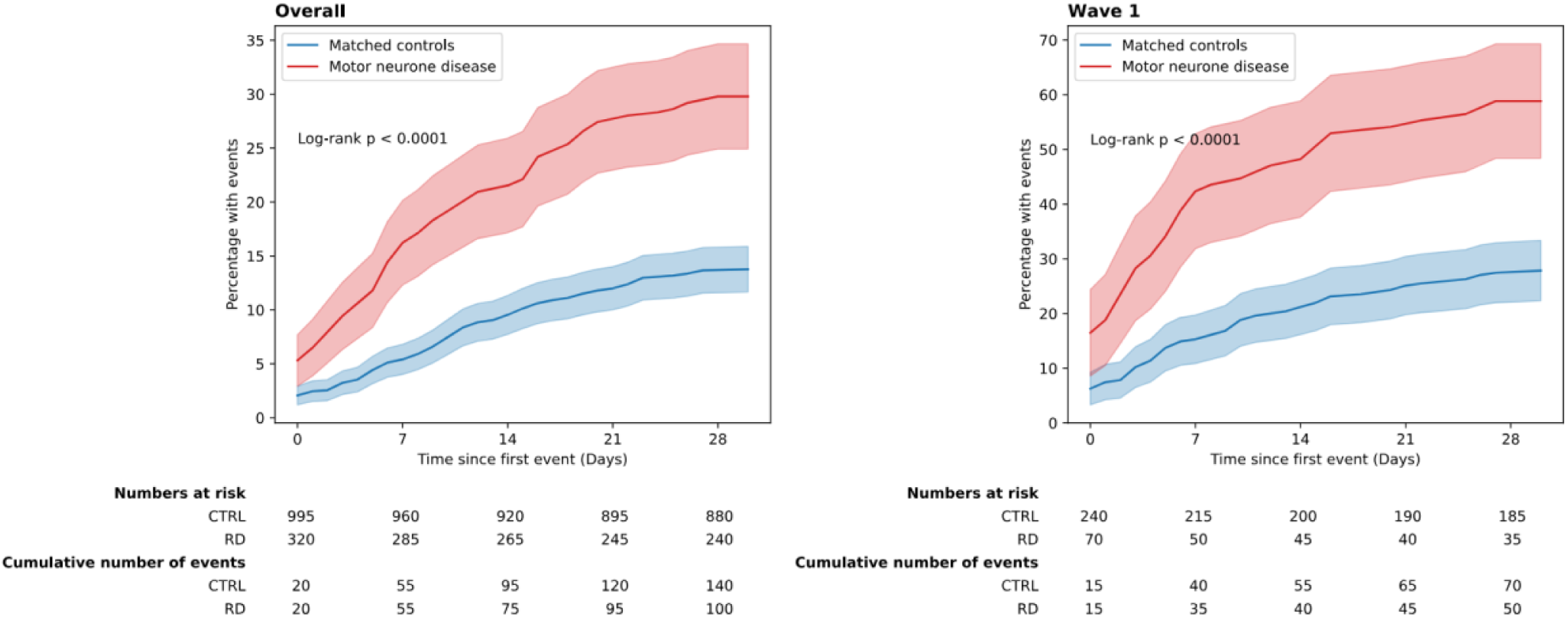
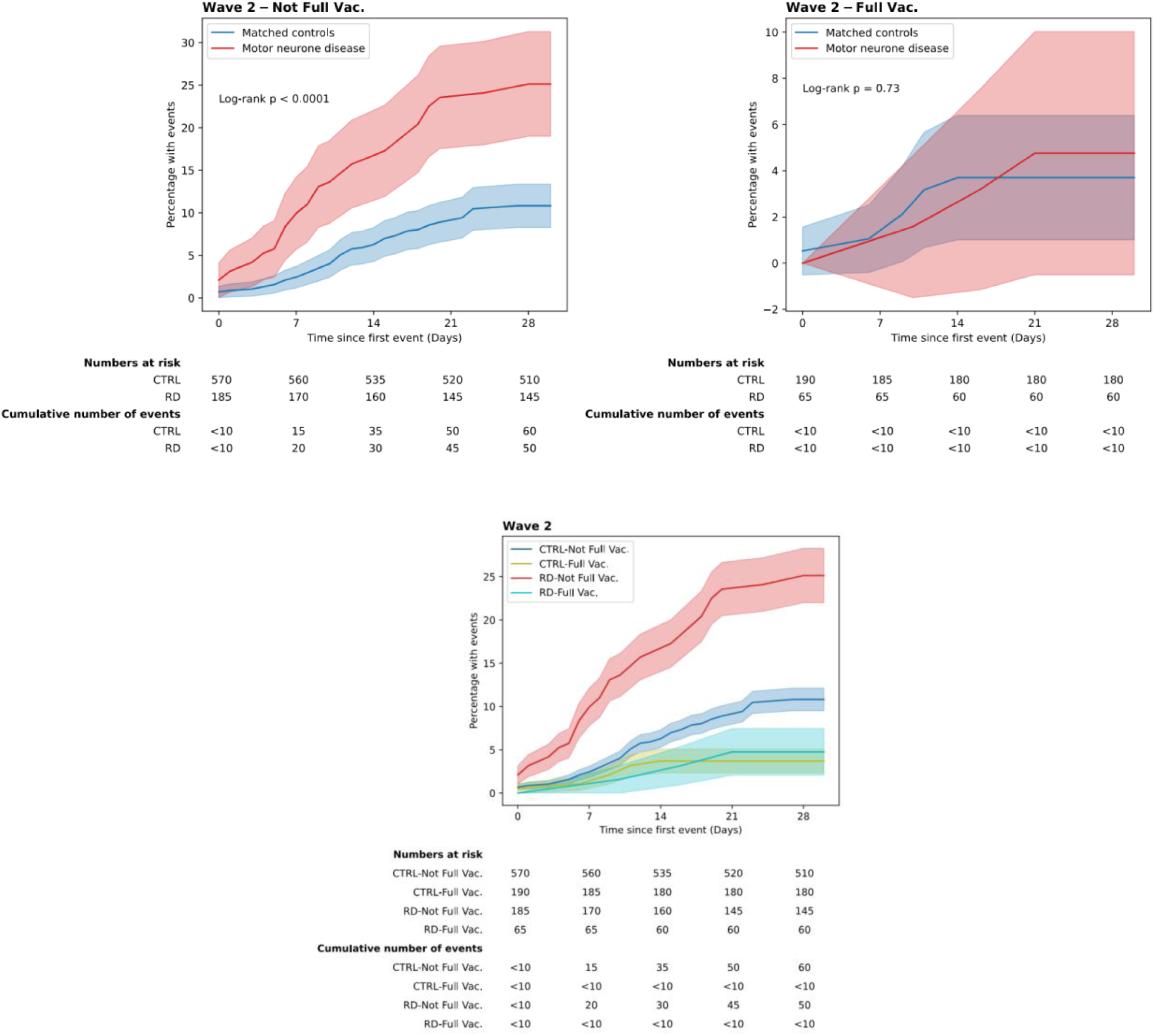
Survival analysis of MND individuals on COVID-19 associated mortality. Kaplan-Meier curves showing the cumulative percentage of events over time (in days) for individuals with motor neurone disease (MND) and matched controls. Left panel (Overall): Survival analysis for the full cohort, demonstrating a significantly higher event rate in the MND group compared to matched controls (log-rank p < 0.0001). Right panel (Wave 2 – Not Fully Vaccinated): Survival analysis for individuals in the second wave of the pandemic who were not fully vaccinated, showing a similar trend of increased risk in the MND group (log-rank p < 0.0001). Shaded regions represent 95% confidence intervals. Below each plot, tables display the number of individuals at risk and cumulative events over time.

During Wave 1, MND patients showed particularly elevated risk (HR=2.97, 95% CI: 1.97-4.48, log-rank p<0.0001). In Wave 2, non-vaccinated MND patients maintained significantly higher risk (HR=2.56, 95% CI: 1.93-3.38, log-rank p<0.0001) compared to matched non-vaccinated controls. However, among fully vaccinated individuals in Wave 2, the mortality difference between MND patients and controls was no longer statistically significant (HR=1.27, 95% CI: 0.69-2.34, log-rank p=0.73), suggesting that vaccination substantially mitigated the excess risk associated with MND.

The cumulative percentage of COVID-19 events remained consistently higher in non-vaccinated MND patients compared to both vaccinated MND patients and control groups. At 28 days post-first COVID-19 event, approximately 25% of non-vaccinated MND patients had experienced a mortality event, compared to approximately 4-5% in the fully vaccinated MND cohort.

Our findings indicate that while MND patients face substantially elevated COVID-19 mortality risk, full vaccination appears to effectively reduce this excess risk to levels comparable with the general population.

## 4. Discussion

As far as we know, this is the first study that is able to use a whole population linked health datasets for studying MND. We were able to derive the largest MND cohort (12,240) during a six-year period from the start of 2014. This made it possible to provide an accurate overall period prevalence of MND in England as well as datasets with both traditional analytics and advanced AI technologies, we carried out a comprehensive analysis on three tasks for MND: prediction (1, 3 and 5 years before coded diagnosis), diagnosis as well as risk assessment of COVID-19 associated mortality.

For MND screening (the prediction tasks), the MNDA guideline (the MND red flag list) was shown to have poor to moderate discrimination (AUROC: 0.62-0.63). It was even worse than the general purpose foundation AI model, GPT-4, on two early diagnosis tasks: predicting MND 1 or 3 years prior to coded diagnosis. The ensemble approach, combining knowledge graph, guideline and real world data (RWD), was able to achieve best discrimination across all tasks, in particular, with an AUROC score of 0.68 for the 5-year prediction.

For diagnosing MND, the MNDA guideline was shown to have a good discrimination power (AUROC=0.703). However, its overall performance assessed by F1 score (a balance between sensitivity and PPV) was low (0.435), which was only slightly better than the GPT-4 model (AUROC: 0.696; F1: 0.380). The ensemble approach achieved significant higher overall performance (28% increase on F1). The RWD model was able to achieve the highest discrimination (AUROC: 0.776).

The missing of predictive phenotypes (early symptoms or diagnosis) seemed to be the main reason for the relatively poor performances of the MNDA guideline. The ICD-10 chapters of the red flag list were well covered by knowledge graph (KG) derived ones, which is understandable as both are supposed to be curated from the scientific evidence. However, KG was shown to identify novel areas including the diseases of the digestive and respiratory systems as well as mental disorders, which was probably the reason for its higher performances than the guideline. The machine learning based RWD model identified seven disease areas that were overlooked by MNDA. Five of those were shown to contain phenotypes having significant effects on both diagnosing and predicting MND.

This study revealed symptoms and comorbidities that are strong predictors of MND including dysphagia, abnormal weight loss, essential hypertension, pure hypercholesterolaemia, unspecified acute lower respiratory infection and epilepsy. Among these, dysphagia^52^ (in the red flag list) and abnormal weight loss^53^ have been widely reported in the literature with high associations with MND. There has been inconsistent evidence on associations of essential hypertension and pure hypercholesterolaemia with MND. In fact, a recent study in Sweden reported protective effects of both conditions for MND.^54^ However, their MND cohort size was 8 times smaller. Our findings also indicated that epilepsy was associated with increased likelihood of MND in both diagnosis and 3-year prediction analysis. We also identified a range of conditions associated with reduced odds of MND: type 2 diabetes^55^ and asthma were with the biggest protective effects followed by chronic kidney disease, Alzheimer’s Disease as well as atrial fibrillation and flutter, peripheral vascular disease and stroke. Prostate cancer and acute sinusitis were particularly shown to be protective in 3-year MND prediction analysis.

A range of computational methods were compared and contrasted in this study for phenotyping model derivation, diagnostic analytics and risk assessments. For phenotyping MND, although the GPT-4 derived models in general performed not great, it did show performances on par with the MNDA red flag list, which was encouraging as it was not adapted for the particular task or on any MND patient data. Knowledge graphs showed great potential in surfacing insights for MND diagnosis with the help of proper ranking algorithms. An interesting finding was that the ‘simple’ graph algorithm of PageRank outperformed more advanced ones like graph Transformers. Real-world data derived models demonstrated superior performances among individual models in all tasks, indicating the great value of hypothesis-free analytics on routinely collected EHRs. For diagnosis and prediction tasks, Random Forest outperformed logistic regression, Support Vector Machine as well as the LSTM model (a type of recurrent neural networks).

Our findings highlight the significantly higher COVID-19 mortality among MND patients during the early pandemic waves, underscoring the urgent need for targeted interventions for this vulnerable group. A separate study in the US conducted equally found that mortality for those living with MND was higher during COVID-19 years compared to pre-COVID-19 years.^6^ However, the sharp decline in COVID-19-related mortality among fully vaccinated individuals reinforces the importance of vaccination in mitigating risks.

While this study presents a novel integration of computational methods with the Red Flag list, several limitations must be acknowledged. First of all, for our study population, it is important to note that this discrepancy is due to a lack of registration data, this data includes temporary registrants and therefore though this study involves the English population, there is a greater number in this study. In addition, it’s important to highlight that primary care data was limited in this study, where many early phenotypes are likely to first appear - and this limits the ability to fully capture the diagnostic window. This exclusion was necessary as SNOMED-CT coverage was incomplete, particularly for signs and symptoms. Additionally, the use of ICD-10 codes introduces potential inaccuracies, as these codes may not fully capture the clinical phenotype displayed by patients due to poor coding practices or lack of financial incentive for their recording. Patients’ symptoms may be incompletely or inaccurately recorded in the notes, and further errors can be introduced when an ICD-10 code is selected for an episode. Equally, as stringent as we have tried to be within this study, there is the risk of not selecting the most accurate ICD code for a particular phenotype from the KG.

The heterogeneity of MND presents a challenge in model performance. The disease has multiple subtypes, including ALS, PBP, PLS, and PMA, each with distinct clinical trajectories. This variation may limit the ability of computational approaches to generalise across the full spectrum of MND presentations. The reliance on retrospective data from the nationwide dataset also introduces biases related to healthcare access and diagnostic delays, which may influence the observed symptom progression patterns.

Finally, while our approach demonstrates the potential for improved diagnostic accuracy through data-driven insights, the feasibility of integrating such methodologies into clinical practice remains an open question. Future work should explore strategies for embedding KG-based tools into electronic health records and real-time decision support systems to Facilitate seamless adoption in clinical settings.

### Contributions

YA, JW and HW conceived and designed the study. YA constructed the subgraph from PrimeKG, produced a phenotype list from the knowledge graph, GPT-4 and clinical guidelines, conducted manual mapping of phenotypes to SNOMEDCT and ICD-10 codes and evaluated phenotype importance in disease. JW conducted NHS patient data curation and statistics, control cohort matching, real-world phenotype modeling, prediction model analysis, and COVID-19 related analyses. YA, JW and HW wrote the first draft. All authors were responsible for writing, reviewing, and editing the manuscript.

## Competing interests

### Ethical and Regulatory Approvals

The data used in this study are available in NHS England’s Secure Data Environment (SDE) service for England, but as restrictions apply they are not publicly available (https://digital.nhs.uk/services/secure-data-environment-service). The CVD-COVID-UK/COVID-IMPACT programme, led by the BHF Data Science Centre (https://bhfdatasciencecentre.org/), received approval to access data in NHS England’s SDE service for England from the Independent Group Advising on the Release of Data (IGARD) (https://digital.nhs.uk/about-nhs-digital/corporate-information-and-documents/independent-group-advising-on-the-release-of-data) via an application made in the Data Access Request Service (DARS) Online system (ref. DARS-NIC-381078-Y9C5K) (https://digital.nhs.uk/services/data-access-request-service-dars/dars-products-and-services). The CVD-COVID-UK/COVID-IMPACT Approvals & Oversight Board (https://bhfdatasciencecentre.org/areas/cvd-covid-uk-covid-impact/) subsequently granted approval to this project to access the data within NHS England’s SDE service for England. The de-identified data used in this study were made available to accredited researchers only. Those wishing to gain access to the data should contact in the first instance.

### Funding

This work was supported by the UK Engineering and Physical Sciences Research Council (EPSRC) [EP/Y035216/1] Centre for Doctoral Training in Data-Driven Health (DRIVE-Health) at King’s College London, with additional support from LifeArc. AAK is funded by The Motor Neurone Disease Association (MNDA), NIHR Maudsley Biomedical Research Centre and ALS Association Milton Safenowitz Research Fellowship, the Darby Rimmer MND Foundation, LifeArc, and the Dementia Consortium. AAK is supported by the UK Dementia Research Institute through UK DRI Ltd, principally funded by the Medical Research Council.

## Data Availability

The data used in this study are available in NHS England’s Secure Data Environment (SDE) service for England, for inquiries about data access, please see www.healthdatagateway.org/dataset/7e5f0247-f033-4f98-aed3-3d7422b9dc6d or

## Appendix

## Supplementary Methods

### Ranking Algorithms

Phenotype importance of phenotypic nodes was ranked by four complementary approaches:

1. **Graph-based**: PageRank and Hyperlink-Induced Topic Search (HITS) (42,43) were used to rank nodes based on graph topology.
2. **Graph Embeddings:** Node2Vec (44) was employed to generate numerical representations (embeddings) of nodes. These embeddings were combined with degree centrality (weight 0.7) and betweenness centrality (weight 0.3) scores.
3. **Graph Machine Learning:** Graph Convolutional Networks (GCNs) (48) and Graph Transformers were utilized to learn complex node representations and predict importance scores.

Kendall’s Tau correlation analysis was used to compare node rankings between MND and Hepatitis subgraphs. A statistically significant correlation was interpreted as indicating an insufficiently discriminatory subgraph size for disease-specific phenotype identification.

Ranked phenotypes from these algorithms were manually mapped to their corresponding ICD-10 codes. Where a direct ICD-10 code was unavailable, a higher-level or best-related ICD-10 code was assigned to ensure comprehensive coverage.

### Using a public Knowledge Graph for evaluation

We used PrimeKG, a KG designed for precision medicine analyses (30). This graph contains information from 20 high-quality resources, spanning 17,080 diseases and more than 4 million relationships, which contain disease-associated protein perturbations, pathways, anatomical and phenotypic scales, and approved drugs with their therapeutic actions.

To focus our analysis on MND, we selected a subgraph that was 2 hops away from the central MND node and removed self-loops and isolated nodes to improve computational efficiency. We decided the subgraph size through the construction of two subgraphs, a MND subgraph and a Hepatitis (control) subgraph both focused on disease-associated phenotypes. All forms of hepatitis were selected as a control for the subgraph comparison because it represents a disease group which has a different pathophysiological mechanism and clinical manifestation compared to MND. The difference allows us to assess whether the structure and relationships within the MND subgraphs are specific to MND. Secondly, Hepatitis is well-represented in knowledge graphs making it suitable for evaluation of subgraph differences. Subgraphs were generated at four levels, with one hop away being referred to as K1 and four hops away being referred to as K4, which represents the increasing distances from the central node. PageRank rankings were calculated for each subgraph level and Kendall Tau correlation analysis was used to compare the similarity of the rankings between the MND and Hepatitis subgraphs. A statistically significant correlation would suggest an inappropriate subgraph size to discriminate between different disease phenotypes.

### MND Association Red Flag List

We used the Red Flag Diagnosis tool, developed by the MND Association in tandem with the RCGP. This tool, designed to speed up referrals to neurology and improve diagnostic accuracy, is tailored to identify early MND indicators in primary care. Using this tool, we obtained a curated list of phenotypes commonly associated with early stage MND based on clinical expertise. Appendix Table 1 shows the 20 phenotypes listed in the Red Flag Diagnosis tool (8,41)

### The evaluation of phenotyping models in MND early diagnosis

We constructed a framework for early MND detection using phenotypes list obtained from KG ranking algorithm, clinical guidelines, and real world data driven approach, as well as demographic data (age, sex, ethnicity). These features were encoded as sparse matrices to manage high dimensionality, with MND diagnosis as the binary target. To address class imbalance, we applied SMOTE on training folds and incorporated class weights during model fitting.

Regarding the predictive modelling, four models were compared:

1. **L1-regularised logistic regression** (C=0.1) for interpretable coefficients.
2. **Random forest** (200 trees, max depth 10) to capture non-linear effects.
3. **Support vector machine** (RBF kernel) to optimise decision boundaries.
4. **LSTM network** (layers 128–64–32, batch normalisation, dropout 0.2) to model temporal dependencies.

### Phenotype Similarity and Statistical Analysis

Phenotype similarity analysis was undertaken to identify similarities between several components, including (a) ranking outputs from the different algorithms compared against each other; (b) ranking outputs from algorithms against clinical guidelines; (c) Ranking outputs from algorithms against Real-World Data (RWD) derived from EHR at three different time points: 1-year, 3 years and 5 years and (d) Clinical Guidelines Comparison to RWD at three different timepoints: -year, 3 years and 5 years.

To assess the concordance between different ranking methodologies, we used Kendall Tau and Jaccard similarity. Kendall Tau evaluates the ordinal agreement between two ranked lists, with values ranging from −1 (complete disagreement) to 1 (perfect agreement). A positive Kendall Tau correlation with statistical significance (p < 0.05) indicates substantial agreement in the ranking order. We applied Kendall Tau to compare rankings generated by different algorithms and to assess the alignment of algorithm-derived rankings with clinical guidelines. Jaccard Similarity measures the overlap between two sets of items, with values ranging from 0 (no overlap) to 1 (complete overlap). We used Jaccard Similarity to quantify the agreement between algorithm-generated rankings and real-world data (RWD) phenotypes at different time points (1, 3, and 5 years), as well as between clinical guidelines and RWD at these time points. Statistically significant Jaccard Similarity scores (p < 0.05) indicate substantial overlap between the compared sets.

### Real World Data enabling phenotype extraction

Phenotypic data was extracted from the national dataset using ICD-10 coding for secondary care using a hierarchical approach. For this study, we focused on secondary care data rather than primary care records for two key reasons. First, the primary care data available in our dataset was sparsely coded and lacked granularity for phenotypic evaluation. Second, secondary care records—typically collected in specialist and hospital settings— may offer more clinical detail and more comprehensive documentation of complex symptom profiles, investigations, and diagnoses. By analysing secondary care data, we aimed to capture a broader and more accurate range of phenotypes associated with MND, many of which may not yet be fully appreciated or recorded in primary care. These insights may inform and refine primary care decision-making, aiding in better recognition and referral of suspected MND cases. This involved mapping ICD codes to broader categories, capturing overarching disease patterns instead of focusing on highly specific diagnoses. When multiple ICD-10 codes fell under the same broad category, the specific ICD-10 code was explicitly listed to retain clinical relevance. A matched control set was constructed by aligning individuals based on age, sex, and ethnicity. This enabled a comparative analysis of the most frequent phenotypes in the control population versus those observed in MND patients.

### Real World Data based phenotype selection

For phenotype identification, we utilized the Human Phenotype Ontology (HPO), specifically focusing on the terms under the branch of “Phenotypic abnormality” (HP:0000118), which yielded xxx distinct terms. These HPO terms were systematically mapped to corresponding ICD-10 and SNOMED-CT codes to identify relevant phenotypes from both GDPPR and HES. We employed multiple feature selection approaches, including LASSO regularisation, logistic regression, correlation analysis, Chi-squared testing, and mutual information criteria, to identify the 20 most relevant phenotypes. The top 20 phenotypes were chosen to ensure a direct comparison to the Red Flag List, which also consists of 20 clinically curated phenotypes.

## Supplementary results

### Phenotype List Similarity Analysis

We compared the correlation heatmap between 4 subgraphs (defined in methods) for ALS and Hepatitis respectively. There is a difference in the rankings and presence of phenotypes for 1-hop out from the central ALS node compared to 1-hop out from central Hepatitis node (K1-ALS and K1-Hepatitis respectively), with a Kendall Tau score of −0.122, indicating little to no similarity in phenotypes in the set as well as the ranking order. However, from K3 onwards, we start to see a statistically significant similarity in PageRank rankings when comparing the two sets of lists. For K3 comparisons, we see a 0.292 score for Kendall Tau, indicating that there is moderate overlap in the rankings produced by the two graphs from two different diseases. At K4, however, we see a greater overlap with a Kendall Tau of 0.793 and this statistically significant result demonstrates that at this subgraph size, there is little to no difference in rankings. This suggested that the differences in the subgraphs were best placed at the K2 size for ALS, and we focused our generation of rankings based on 2 hops out from the central ALS node.

The ranking produced by the PageRank algorithm has a Jacquard similarity score of 0.329 with the ranking produced by the Graph Embedding method. This, in addition to a statistically significant p-value (p < 0.05) reveals that there is a moderate overlap between the two rankings. Similarly, we see a statistically significant moderate correlation between the GCN ranking outputs and Graph Transformer, with a score of 0.301. All other similarity scores are between −0.145 and 0.131, and are not statistically significant. This further testifies to the diversity in the rankings produced by different algorithms.

When comparing the ranking output to the clinical guidelines, we see that the Jaccard similarity score ranges between 0 to 0.087, indicating that there is virtually no similarity between the two sets of lists. The fact that the highest similarity score barely exceeds 0.08 highlights the lack of significant overlap between each set. This outcome was unexpected, as we would expect the top-ranked phenotypes from each algorithm to show a higher degree of overlap with the guidelines. This low similarity score may indicate that the algorithms prioritised different features in the Knowledge Graph compared to traditional clinical methods of determining important phenotypes

Jaccard similarity analysis of the top 20 most frequent phenotypes at 1-, 3-, and 5-years before MND diagnosis revealed a high degree of overlap across time points. The greatest similarity was observed between the 1-year and 3-year pre-diagnosis periods (0.895) (p < 0.05) indicating that symptoms present at three years before diagnosis are highly consistent of those found closer to diagnosis.

To assess the alignment between MNDA Red Flag List and phenotypes seen in the real world, we performed Jaccard Similarity with the phenotypes from the guidelines and the 20 most frequent phenotypes observed in the dataset at 1-, 3- and 5-year time points. We found that there was no similarity at any time point, with scores consistently at 0.

### Phenotype frequency in the study population

We analysed the most frequent phenotypes for individuals with MND at three predefined time points: up to five years before diagnosis, three years before diagnosis, one year before diagnosis, and across all available years before diagnosis. This allowed us to capture early indicators of MND and examine how phenotypic presentation evolves over time.

When considering all years before diagnosis, clear distinctions emerged between MND patients and control individuals. Several phenotypes were more commonly observed in those with MND, providing strong evidence of their association with the disease. These included aphagia and dysphagia, respiratory failure, gait and mobility abnormalities, speech disturbances, and disorders of the urinary system.

Five years prior to diagnosis, fewer distinct indicators were observed; however, some differences remained. Cataracts were notably more frequent among MND patients, while conditions such as chronic obstructive pulmonary disease (COPD) and nicotine dependence appeared more commonly in the control population.

At three years before diagnosis, phenotypic overlap between MND and control populations remained. Several features such as gastro-oesophageal reflux disease (GORD) and cataracts became increasingly prevalent in the MND group, whereas COPD and malignant neoplasms remained more frequent in the control group.

One year before diagnosis, there was still substantial overlap in commonly occurring conditions between the two populations. However, phenotypes such as GORD, cataracts, and symptoms involving the nervous and musculoskeletal systems were more frequent in the MND group. In contrast, COPD, pneumonia, and nicotine dependence were more commonly found in the control population.

### Phenotype frequency, non-motor symptoms and the MND Association Red Flag List

Urinary symptoms and functional intestinal issues were frequently identified across all time points before diagnosis. In a study by Shojaie et al. (46), non-motor symptoms in ALS were examined through a 20-item questionnaire. Interestingly, the most commonly reported non motor symptom was urinary urgency, a phenotype that we see at a national level but not present within the Red Flag list, as seen in Figure S9. Interestingly, it has been noted within the development of the Red Flag List that these symptoms do not support an MND diagnosis. Equally, constipation was a reported symptom in this questionnaire, relating to functional intestinal issues, a symptom we see at national level. This gastrointestinal symptom in ALS has also been explored in a study which found that pTDP-43 aggregates were found in multiple cell types in the GI tract (47). These findings suggest that autonomic dysfunction may play a larger role in disease progression than previously recognised. Interestingly, the study by Mei et al. (8) also found similar patterns of autonomic dysfunction in MND patients at the primary care level, reinforcing the relevance of these symptoms in disease progression. Given the increasing awareness of non-motor symptoms in neurodegenerative diseases, further research is needed to determine whether these symptoms could serve as early indicators or even predictive markers of MND. The recognition of these symptoms in clinical practice may allow for a more comprehensive understanding of disease onset and a broader framework for early detection.

By incorporating these insights into clinical frameworks, future research should focus on refining diagnostic pathways to include non-motor and early neuromuscular signs, potentially improving early detection and patient outcomes. Expanding this computational approach to other neurodegenerative diseases may reveal shared characteristics in disease progression and symptomatology. Further research is also needed to investigate the role of environmental and lifestyle factors in MND development through large-scale data-driven methods.

### Important Phenotypes for Predicting and Diagnosing MND

Figure S9 gives the important phenotypes derived from the best performing models for both MND diagnosis and 3-year prediction. The subfigure Figure S9a compares and contrasts the phenotypes at ICD-10 chapter level between individual models (knowledge graph, the MNDA guideline and real world data) in the ensemble approach. For the MND diagnosis task (the upper part of Figure 2a), all three individual models used chapter XVIII (Symptoms, signs and abnormal clinical and laboratory findings, not elsewhere classified) and VI (Diseases of the nervous system) with the former being ranked very high in all models. The MNDA guideline and knowledge graph approaches had more commonalities (chapters XVIII, XIII, VI and XXI). The RWD model used five extra chapters with IV (Endocrine, nutritional and metabolic diseases) and IX (Diseases of the circulatory system) being the second and third most important in its ranked list, respectively. For the 3 year MND prediction task (the lower part of Figure 2a), similarly, the knowledge graph model used all four chapters of the MNDA guideline with very similar rankings. The RWD model identified chapters XVIII, IX, V, IV and VI as the most predictive for the MND early diagnosis (3 years).

To investigate the importance of individual phenotypes, the RWD model (via feature selection on the real-world MND cohorts) was used for the analysis because it was the best individual phenotyping model for all diagnosis and prediction tasks. Figure S9b is the forest plot of odds ratios of phenotypes for diagnosing MND derived from a logistic regression model with AUROC=0.742. Phenotypes are grouped into boxes based on their ICD-10 chapters. Those with yellow background highlight phenotype categories that are not included in the MNDA guideline. Having Dysphagia (difficulty swallowing) and abnormal weight loss are associated with the highest risks of MND, followed by Essential hypertension and Unspecified acute lower respiratory infection. Type 2 diabetes mellitus and asthma are the most protective conditions, followed by Chronic kidney disease (stage 3) and Alzheimer disease. All phenotypes except hypertension in chapter IX (Diseases of the circulatory system) demonstrate protective effects.

For the 3-year early diagnosis task, Figure S9c shows the phenotype importance using SHAP method for all phenotypes as well as three demographic features of age, sex and ethnicity. The list is ordered by the impact of features. Age is the most predictive, which, however, has a complex effect pattern as shown in the dependency plot on the top right. For the rage from 60 to 90, age is shown to be associated with higher risks, while other ranges show protective effects. High risk phenotypes include Essential hypertension, Pure hypercholesterolaemia, Dysphagia and Epilepsy, unspecified. Type 2 diabetes mellitus and Asthma show most protective effects. For ethnicity, the bottom right subfigure shows that being in a minority group has a protective effect.

This study revealed symptoms and comorbidities that are strong predictors of MND including dysphagia, abnormal weight loss, essential hypertension, pure hypercholesterolaemia, unspecified acute lower respiratory infection and epilepsy. Among these, dysphagia52 (in the red flag list) and abnormal weight loss53 have been widely reported in the literature with high associations with MND. There has been inconsistent evidence on associations of essential hypertension and pure hypercholesterolaemia with MND. In fact, a recent study in Sweden reported protective effects of both conditions for MND.54 However, their MND cohort size was 8 times smaller. Our findings also indicated that epilepsy was associated with increased likelihood of MND in both diagnosis and 3-year prediction analysis. We also identified a range of conditions associated with reduced odds of MND: type 2 diabetes55 and asthma were with the biggest protective effects followed by chronic kidney disease, Alzheimer’s Disease as well as atrial fibrillation and flutter, peripheral vascular disease and stroke. Prostate cancer and acute sinusitis were particularly shown to be protective in 3-year MND prediction analysis.

## Supplementary Tables

**Table S1:** Top 20 phenotypes related to MND from the MNDA red flag list.

| MNDA red flag list phenotypes |  |
| --- | --- |
| 1. Dysphagia | 11. Shortness of breath on exertion |
| 2. Dysarthria | 12. Excessive daytime sleepiness |
| 3. Tongue fasciculations | 13. Fatigue |
| 4. Focal weakness | 14. Early morning headache |
| 5. Distal weakness | 15. Orthopnoea |
| 6. Falls | 16. Behavioural change |
| 7. Loss of dexterity | 17. Emotional lability |
| 8. Muscle wasting | 18. No sensory features |
| 9. Muscle twitching / fasciculations | 19. Hard to explain respiratory symptoms |
| 10. Cramps | 20. Frontotemporal dementia |

**Table S2:** Jaccard similarity scores comparing phenotype overlap between MNDA red flag list and patient records at 1, 3, and 5 years before MND diagnosis.

| Comparator 1 | Comparator 2 | Jaccard Similarity |
| --- | --- | --- |
| 1-year before diagnosis | MNDA Red Flag List | 0 |
| 3-years before diagnosis | MNDA Red Flag List | 0 |
| 5-years before diagnosis | MNDA Red Flag List | 0 |

**Table S3:** Top 20 phenotypes related to MND from the PrimeKG ranked using the different ranking algorithms.

| Phenotype | PageRank | HITS | GCN | GTrans | GEmb | GPT-4 |
| --- | --- | --- | --- | --- | --- | --- |
| Phenotype 1 | Fatigue | Sensory impairment | Peripheral axonal neuropathy | Distal muscle weakness | Fatigue | Muscle weakness |
| Phenotype 2 | Pain | Dissociated sensory loss | Semantic dementia | Proximal muscle weakness | Dyspnea | Muscle fasciculations |
| Phenotype 3 | Dyspnea | Peripheral neuropathy | Persistent repetition of actions | Apathy | Pain | Muscle cramps |
| Phenotype 4 | Anxiety | Dementia | Persistent repetition of words | Paralysis | Anxiety | Spasticity |
| Phenotype 5 | Muscle Weakness | Frontal lobe dementia | Dysphonia | Difficulty walking | Muscle weakness | Dysarthria |
| Phenotype 6 | Dysphagia | Frontotemporal dementia | Impulsivity | Nausea and vomiting | Dysphagia | Dysphagia |
| Phenotype 7 | Xerostomia | Subcortical dementia | Risk taking | Respiratory failure | Xerostomia | Limb weakness |
| Phenotype 8 | Dysarthria | Cognitive impairment | Primitive reflex | Emotional lability | Agitation | Atrophy of muscle |
| Phenotype 9 | Agitation | Memory impairment | Irritability | Pes cavus | Dysarthria | Hyperreflexia |
| Phenotype 10 | Spasticity | Bradyphrenia | Inappropriate laughter | Areflexia | Gait disturbance | Emotional lability |
| Phenotype 11 | Gait disturbance | Hyporeflexia | Inappropriate sexual behaviour | Gait disturbance | Spasticity | Cognitive impairment |
| Phenotype 12 | Sensory impairment | Gait disturbance | Anomia | Depressivity | Sensory impairment | Depression |
| Phenotype 13 | Cognitive impairment | Spastic gait | Alexia | Hyporeflexia | Cognitive impairment | Sialorrhea |
| Phenotype 14 | Respiratory failure | Unsteady gait | Abulia | Drooling | Emotional lability | Pseudobulbar affect |
| Phenotype 15 | Emotional lability | Difficulty walking | Inflammatory myopathy | Muscle weakness | Drooling | Fatigue |
| Phenotype 16 | Drooling | Shuffling gait | Vocal cord paresis | Sensory impairment | Dementia | Ankle clonus |
| Phenotype 17 | Hyporeflexia | Waddling gait | Weak voice | Cognitive impairment | Hyporeflexia | Gait abnormality |
| Phenotype 18 | Depressivity | Falls | Abnormal breath sound | Dysarthria | Depressivity | Breathing difficulty |
| Phenotype 19 | Dementia | Steppage gait | Airway obstruction | Agitation | Areflexia | Weight loss |
| Phenotype 20 | Areflexia | Gait apraxia | Sneeze | Xerostomia | Difficulty walking | Sleep disturbance |

**Table S4:** Performance comparison of random forest models using MNDA red flag list, graph-based features from PrimeKG, and HPO-driven real-world data 1 year before MND diagnosis.

| Method | Model | Time Period | Accuracy | ROC AUC | Precision | Recall | F1 |
| --- | --- | --- | --- | --- | --- | --- | --- |
| guideline_graph_ensemble | Random Forest | 1y | 0.51 | 0.61 | 0.31 | 0.78 | 0.44 |
| **guideline_graph_hpo_ensemble** | **Random Forest** | **1y** | **0.61** | **0.66** | **0.36** | **0.72** | **0.48** |
| guideline_hpo_ensemble | Random Forest | 1y | 0.6 | 0.65 | 0.35 | 0.73 | 0.47 |
| hpo_chi | Random Forest | 1y | 0.57 | 0.64 | 0.34 | 0.74 | 0.46 |
| hpo_corr | Random Forest | 1y | 0.59 | 0.65 | 0.35 | 0.73 | 0.47 |
| hpo_lasso | Random Forest | 1y | 0.59 | 0.65 | 0.35 | 0.74 | 0.47 |
| hpo_log | Random Forest | 1y | 0.59 | 0.65 | 0.35 | 0.74 | 0.47 |
| hpo_mi | Random Forest | 1y | 0.59 | 0.65 | 0.35 | 0.74 | 0.47 |
| gcn_top20 | Random Forest | 1y | 0.48 | 0.6 | 0.29 | 0.77 | 0.42 |
| gemb_top20 | Random Forest | 1y | 0.51 | 0.63 | 0.31 | 0.76 | 0.44 |
| gpt_top20 | Random Forest | 1y | 0.51 | 0.64 | 0.31 | 0.78 | 0.44 |
| guideline_top20 | Random Forest | 1y | 0.5 | 0.62 | 0.3 | 0.76 | 0.43 |
| hits_top20 | Random Forest | 1y | 0.5 | 0.61 | 0.3 | 0.75 | 0.42 |
| pagerank_top20 | Random Forest | 1y | 0.51 | 0.63 | 0.31 | 0.76 | 0.44 |
| transformer_top20 | Random Forest | 1y | 0.49 | 0.61 | 0.3 | 0.76 | 0.43 |

The performance of models is evaluated using standard classification metrics: area under the receiver operating characteristic curve (ROC AUC), F1 score, precision, recall, and accuracy. Confusion matrix elements are also reported, including true negatives (TN), false positives (FP), false negatives (FN), and true positives (TP).

**Table S5.** Performance comparison of random forest models using MNDA red flag list, graph-based features from PrimeKG, and HPO-driven real-world data 3 years before MND diagnosis.

| Method | Model | Time Period | Accuracy | ROC AUC | Precision | Recall | F1 |
| --- | --- | --- | --- | --- | --- | --- | --- |
| guideline_graph_ensemble | Random Forest | 3y | 0.52 | 0.61 | 0.31 | 0.78 | 0.45 |
| **guideline_graph_hpo_ensemble** | **Random Forest** | **3y** | **0.61** | **0.67** | **0.37** | **0.75** | **0.49** |
| guideline_hpo_ensemble | Random Forest | 3y | 0.6 | 0.66 | 0.36 | 0.75 | 0.48 |
| hpo_chi | Random Forest | 3y | 0.58 | 0.65 | 0.34 | 0.75 | 0.47 |
| hpo_corr | Random Forest | 3y | 0.6 | 0.66 | 0.36 | 0.74 | 0.48 |
| hpo_lasso | Random Forest | 3y | 0.59 | 0.65 | 0.35 | 0.74 | 0.48 |
| hpo_log | Random Forest | 3y | 0.6 | 0.65 | 0.35 | 0.72 | 0.48 |
| hpo_mi | Random Forest | 3y | 0.6 | 0.65 | 0.35 | 0.74 | 0.48 |
| gcn_top20 | Random Forest | 3y | 0.48 | 0.6 | 0.29 | 0.78 | 0.43 |
| gemb_top20 | Random Forest | 3y | 0.52 | 0.63 | 0.31 | 0.76 | 0.44 |
| gpt_top20 | Random Forest | 3y | 0.51 | 0.63 | 0.31 | 0.77 | 0.44 |
| guideline_top20 | Random Forest | 3y | 0.5 | 0.62 | 0.3 | 0.77 | 0.43 |
| hits_top20 | Random Forest | 3y | 0.5 | 0.61 | 0.3 | 0.76 | 0.43 |
| pagerank_top20 | Random Forest | 3y | 0.52 | 0.63 | 0.31 | 0.76 | 0.44 |
| transformer_top20 | Random Forest | 3y | 0.49 | 0.61 | 0.3 | 0.78 | 0.43 |

The performance of models is evaluated using standard classification metrics: area under the receiver operating characteristic curve (ROC AUC), F1 score, precision, recall, and accuracy. Confusion matrix elements are also reported, including true negatives (TN), false positives (FP), false negatives (FN), and true positives (TP).

**Table S6.** Performance comparison of random forest models using MNDA red flag list, graph-based features from PrimeKG, and HPO-driven real-world data 5 years before MND diagnosis.

| Method | Model | Time Period | Accuracy | ROC AUC | Precision | Recall | F1 |
| --- | --- | --- | --- | --- | --- | --- | --- |
| guideline_graph_ensemble | Random Forest | 5y | 0.55 | 0.63 | 0.33 | 0.77 | 0.46 |
| **guideline_graph_hpo_ensemble** | **Random Forest** | **5y** | **0.62** | **0.68** | **0.38** | **0.76** | **0.5** |
| guideline_hpo_ensemble | Random Forest | 5y | 0.61 | 0.67 | 0.37 | 0.77 | 0.5 |
| hpo_chi | Random Forest | 5y | 0.57 | 0.64 | 0.34 | 0.75 | 0.47 |
| hpo_corr | Random Forest | 5y | 0.59 | 0.65 | 0.35 | 0.74 | 0.47 |
| hpo_lasso | Random Forest | 5y | 0.59 | 0.64 | 0.35 | 0.73 | 0.47 |
| hpo_log | Random Forest | 5y | 0.59 | 0.64 | 0.34 | 0.73 | 0.47 |
| hpo_mi | Random Forest | 5y | 0.59 | 0.64 | 0.35 | 0.73 | 0.47 |
| gcn_top20 | Random Forest | 5y | 0.48 | 0.61 | 0.3 | 0.79 | 0.43 |
| gemb_top20 | Random Forest | 5y | 0.52 | 0.63 | 0.31 | 0.76 | 0.44 |
| gpt_top20 | Random Forest | 5y | 0.52 | 0.63 | 0.31 | 0.75 | 0.44 |
| guideline_top20 | Random Forest | 5y | 0.5 | 0.63 | 0.31 | 0.79 | 0.44 |
| hits_top20 | Random Forest | 5y | 0.5 | 0.62 | 0.31 | 0.79 | 0.44 |
| pagerank_top20 | Random Forest | 5y | 0.52 | 0.64 | 0.31 | 0.78 | 0.45 |
| transformer_top20 | Random Forest | 5y | 0.49 | 0.61 | 0.3 | 0.78 | 0.43 |

The performance of models is evaluated using standard classification metrics: area under the receiver operating characteristic curve (ROC AUC), F1 score, precision, recall, and accuracy. Confusion matrix elements are also reported, including true negatives (TN), false positives (FP), false negatives (FN), and true positives (TP).

**Table S7.** Performance comparison of random forest models using MNDA red flag list, graph-based features from PrimeKG, and HPO-driven real-world data all years before MND diagnosis.

| Method | Model | Time Period | Accuracy | ROC AUC | Precision | Recall | F1 |
| --- | --- | --- | --- | --- | --- | --- | --- |
| guideline_graph_ensemble | Random Forest | all | 0.79 | 0.69 | 0.6 | 0.46 | 0.52 |
| guideline_graph_hpo_ensemble | Random Forest | all | 0.74 | 0.73 | 0.49 | 0.65 | 0.56 |
| guideline_hpo_ensemble | Random Forest | all | 0.72 | 0.71 | 0.46 | 0.64 | 0.53 |
| hpo_chi | Random Forest | all | 0.73 | 0.71 | 0.47 | 0.65 | 0.54 |
| hpo_corr | Random Forest | all | 0.74 | 0.72 | 0.48 | 0.64 | 0.55 |
| hpo_lasso | Random Forest | all | 0.74 | 0.72 | 0.48 | 0.65 | 0.55 |
| hpo_log | Random Forest | all | 0.74 | 0.72 | 0.48 | 0.65 | 0.55 |
| hpo_mi | Random Forest | all | 0.74 | 0.71 | 0.48 | 0.63 | 0.54 |
| gcn_top20 | Random Forest | all | 0.5 | 0.63 | 0.3 | 0.76 | 0.43 |
| gemb_top20 | Random Forest | all | 0.58 | 0.68 | 0.34 | 0.69 | 0.45 |
| gpt_top20 | Random Forest | all | 0.77 | 0.7 | 0.58 | 0.28 | 0.38 |
| guideline_top20 | Random Forest | all | 0.78 | 0.7 | 0.58 | 0.35 | 0.43 |
| hits_top20 | Random Forest | all | 0.77 | 0.68 | 0.6 | 0.28 | 0.38 |
| pagerank_top20 | Random Forest | all | 0.77 | 0.7 | 0.59 | 0.31 | 0.4 |
| transformer_top20 | Random Forest | all | 0.78 | 0.68 | 0.72 | 0.22 | 0.34 |

The performance of models is evaluated using standard classification metrics: area under the receiver operating characteristic curve (ROC AUC), F1 score, precision, recall, and accuracy. Confusion matrix elements are also reported, including true negatives (TN), false positives (FP), false negatives (FN), and true positives (TP).

## Supplementary Figures

**Figure S1:**
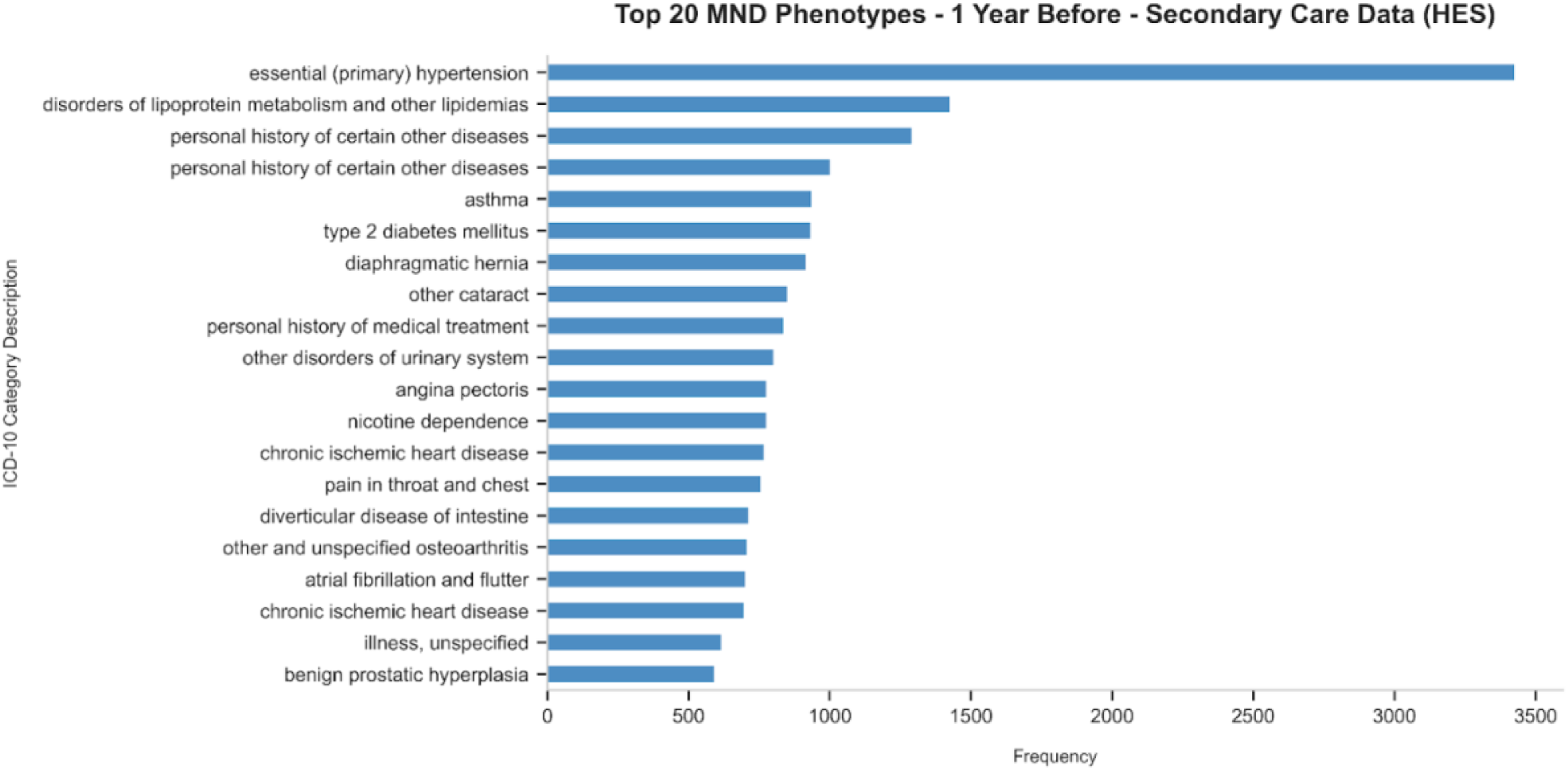
Top 20 phenotypes 1 year before MND diagnosis.

**Figure S2:**
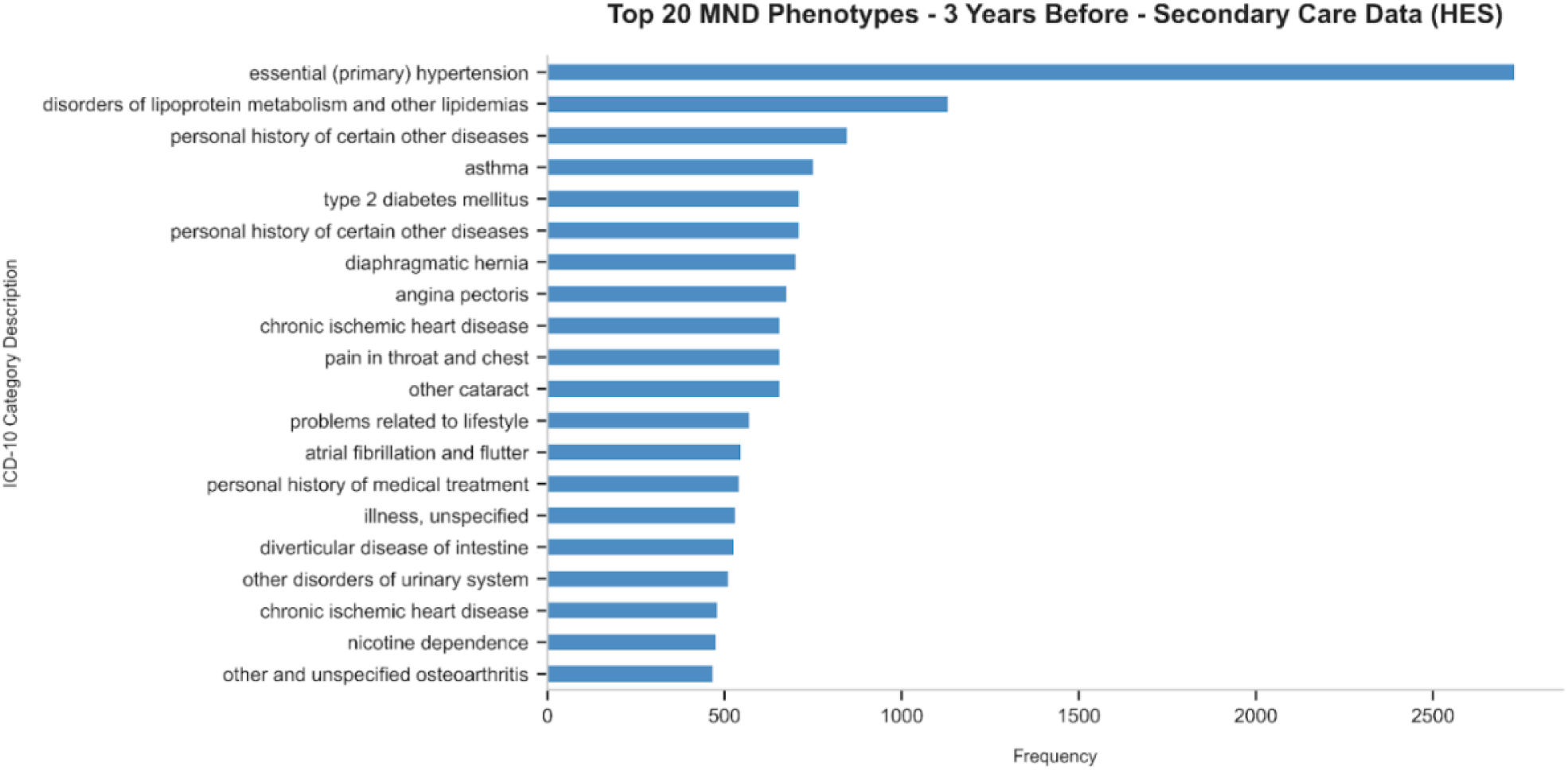
Top 20 phenotypes 3 years before MND diagnosis.

**Figure S3:**
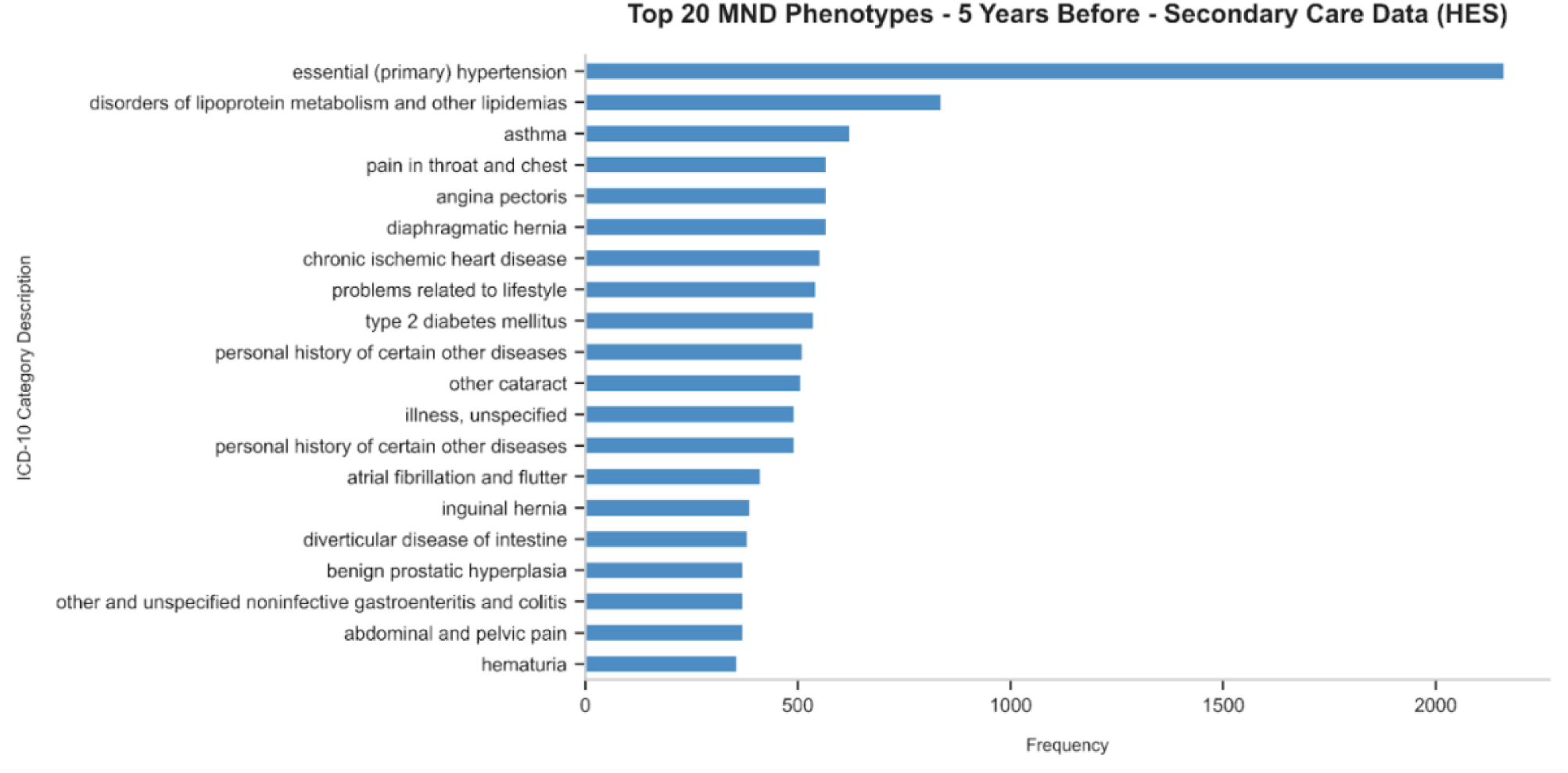
Top 20 phenotypes 5 years before MND diagnosis.

**Figure S4:**
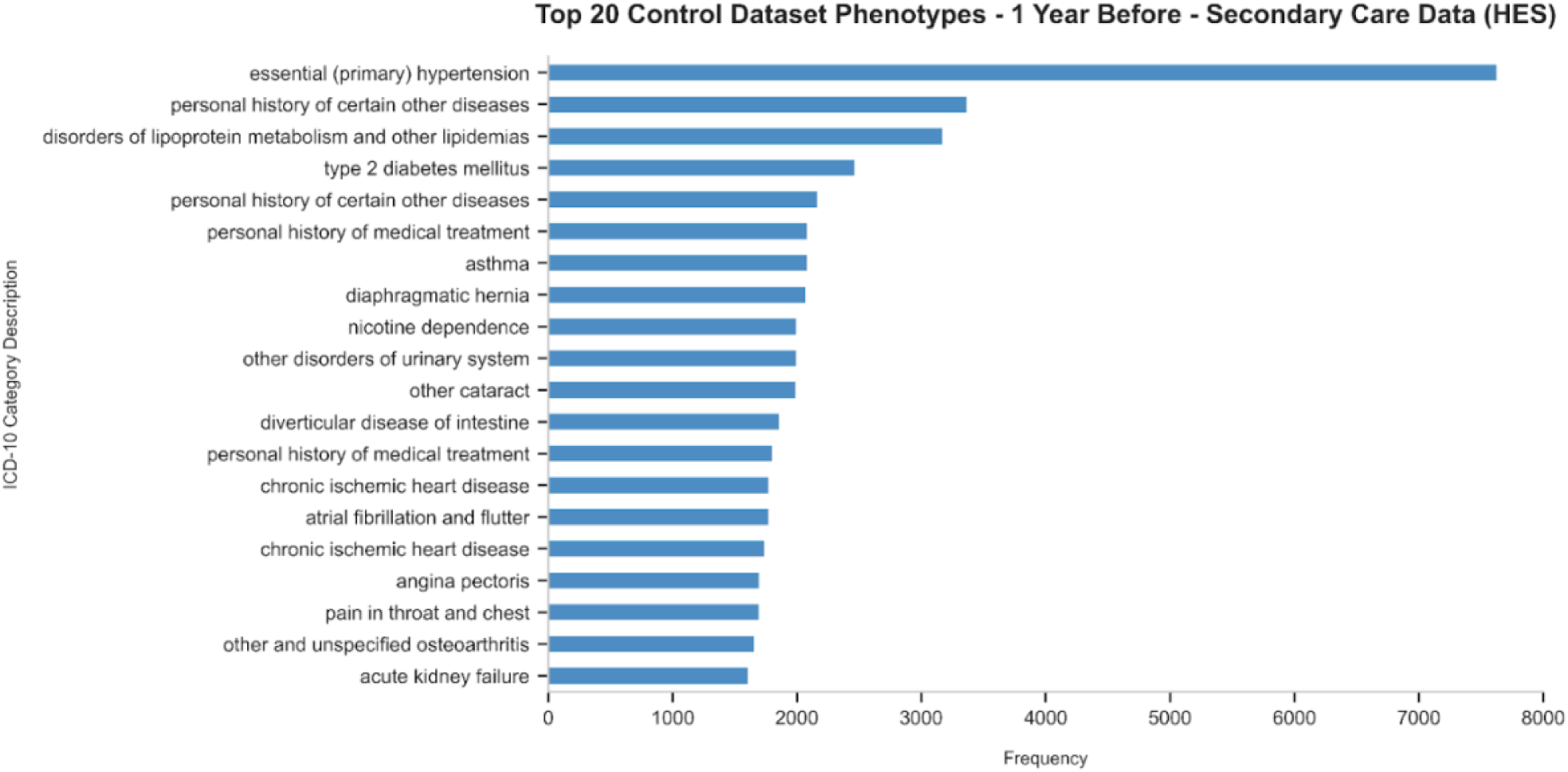
Top 20 phenotypes 1 years among control individuals 1 year before diagnosis.

**Figure S5:**
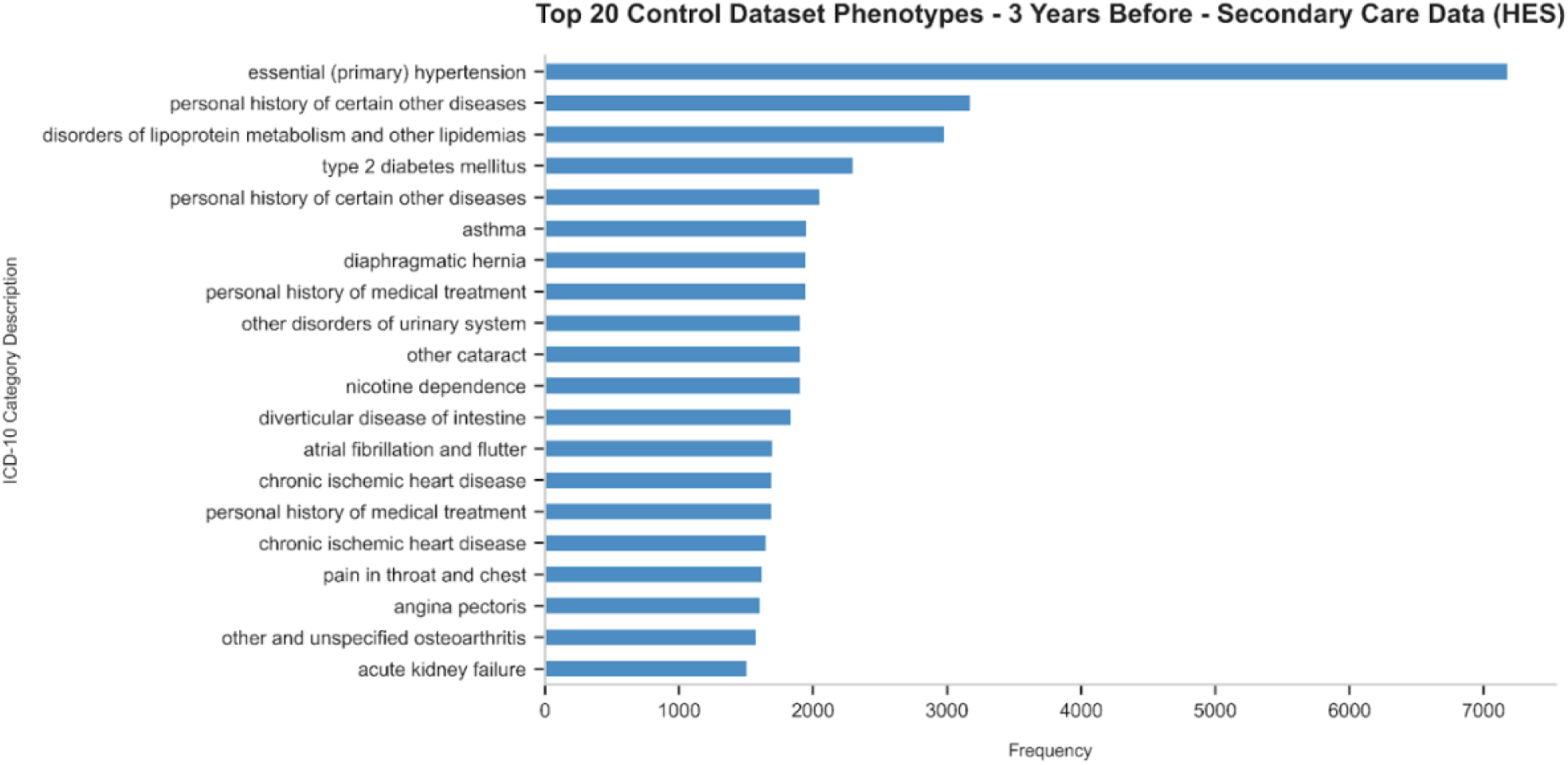
Top 20 phenotypes 3 years among control individuals 3 years before diagnosis.

**Figure S6:**
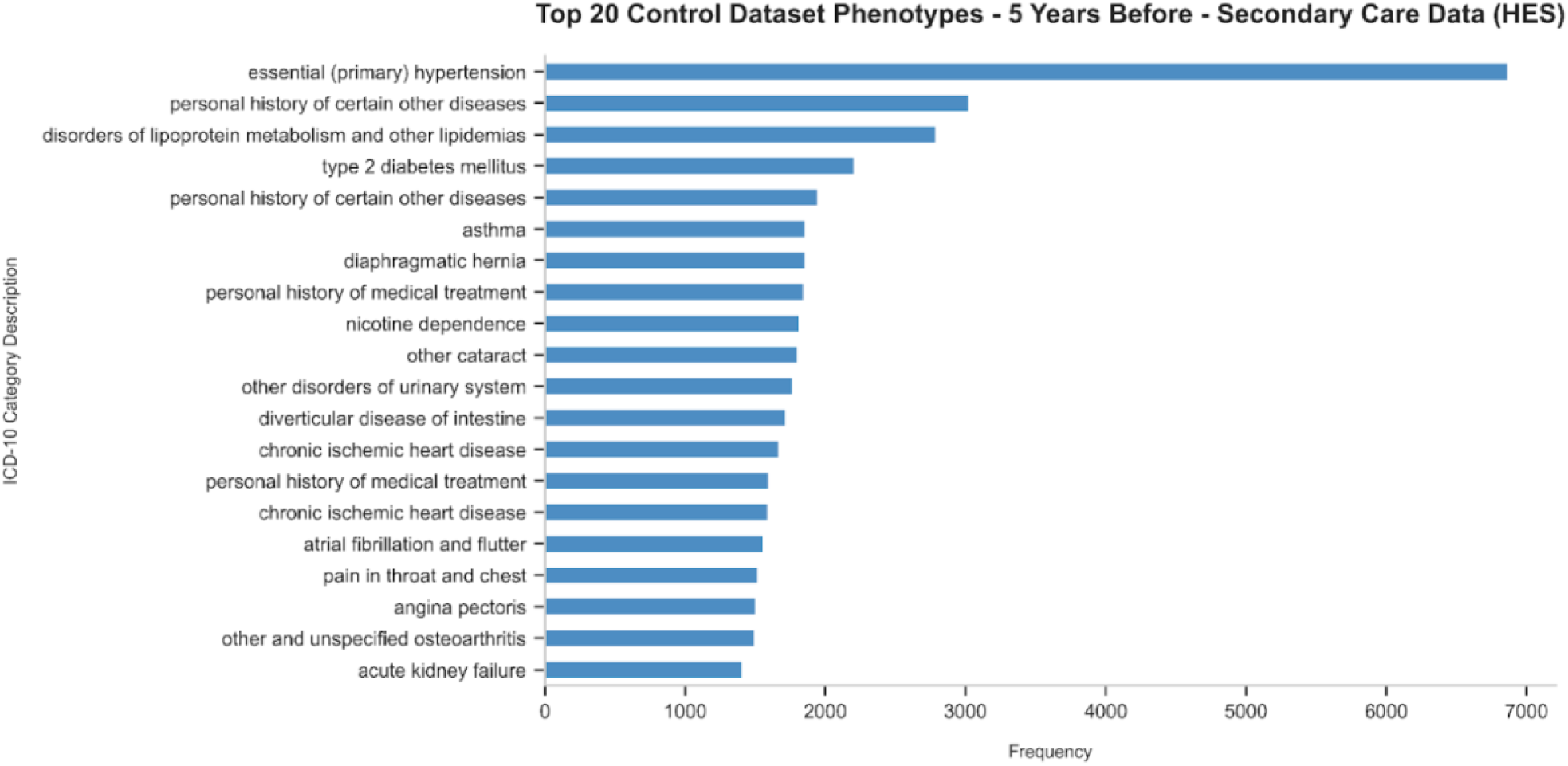
Top 20 phenotypes 5 years among control individuals 5 years before diagnosis.

**Figure S7:**
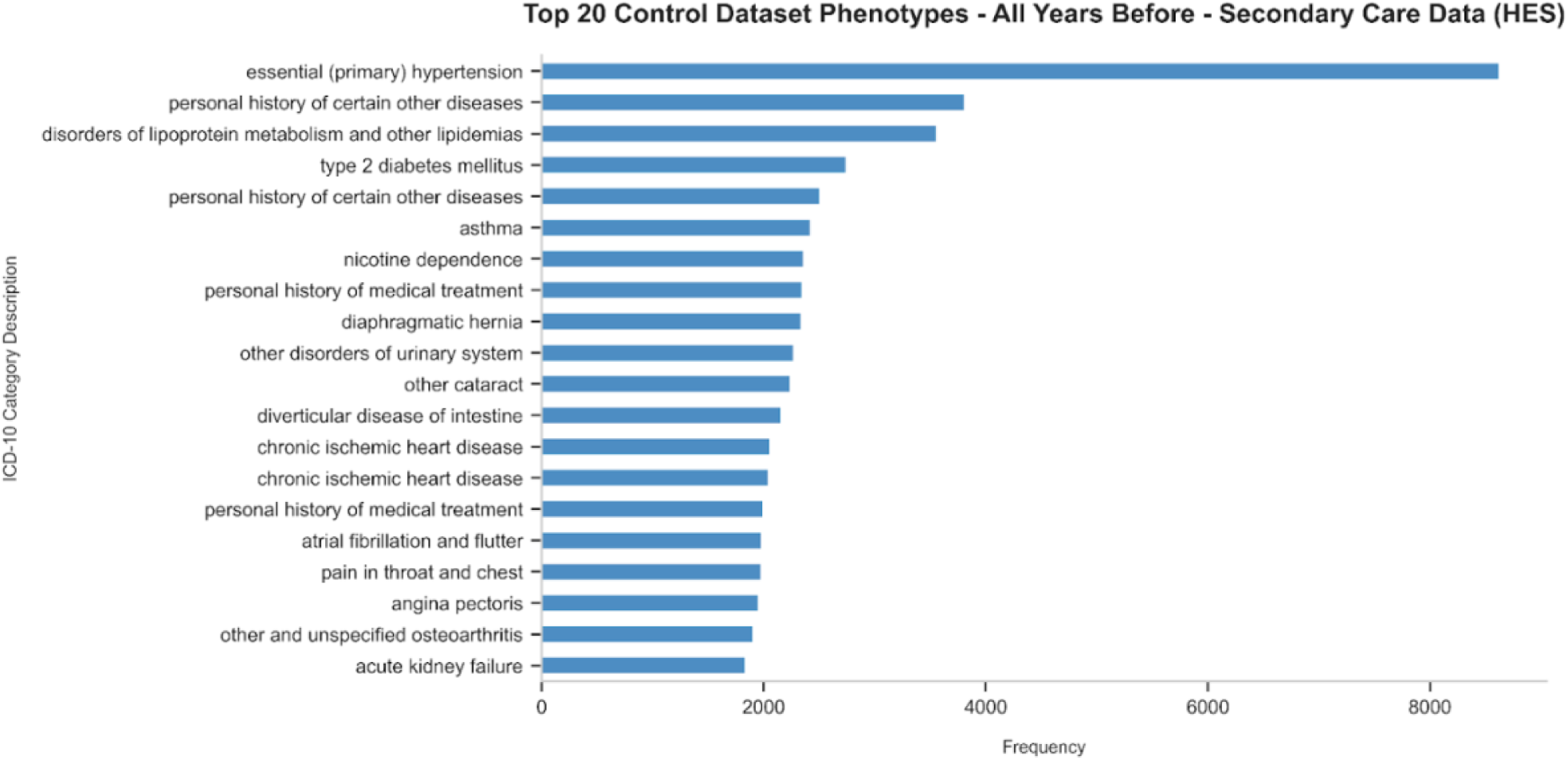
Top 20 phenotypes all years among control individuals all years before diagnosis.

**Figure S8.**
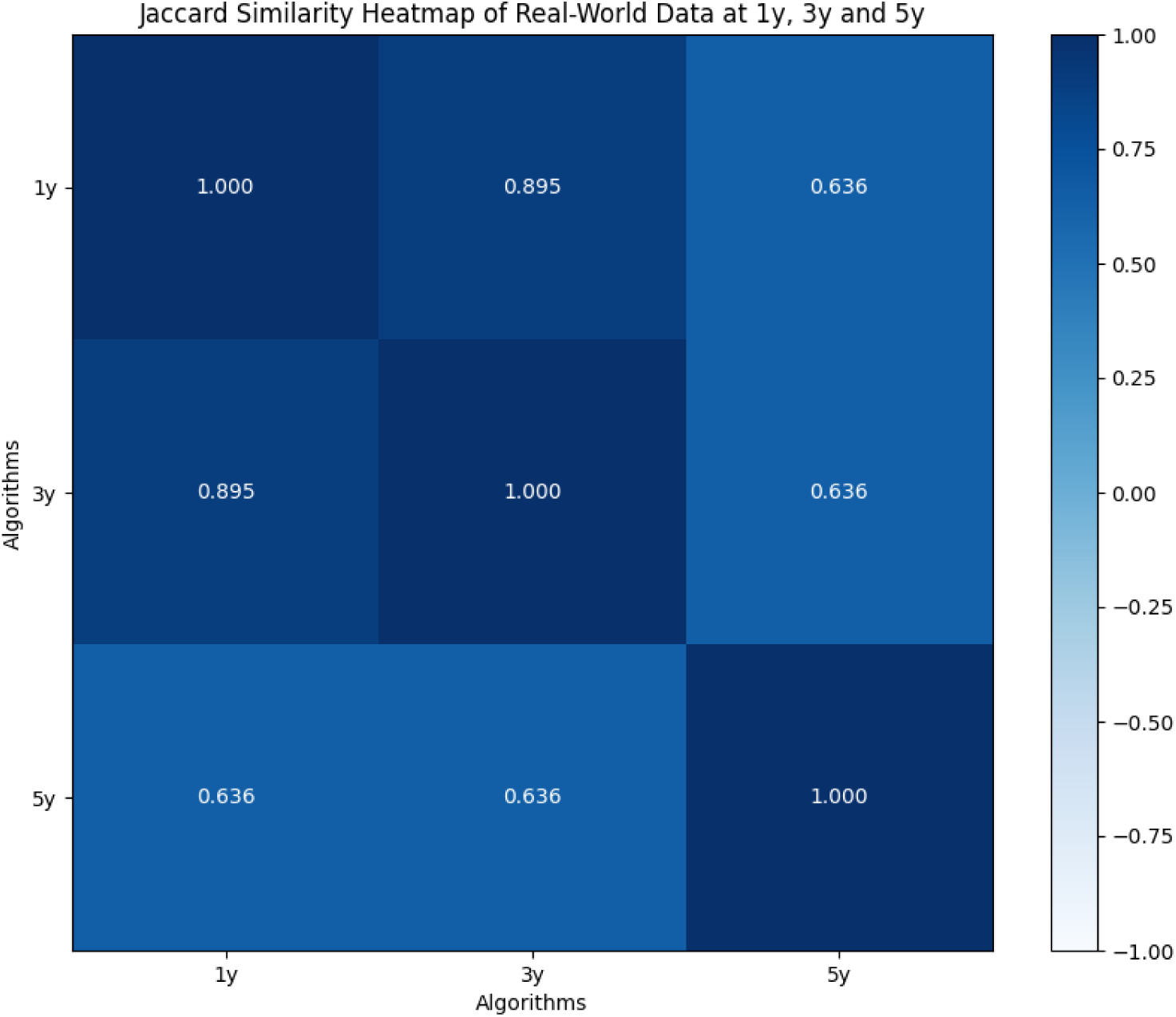
Jaccard similarity heatmap of phenotypes at 1, 3, and 5 years before MND diagnosis.

**Figure S9:**
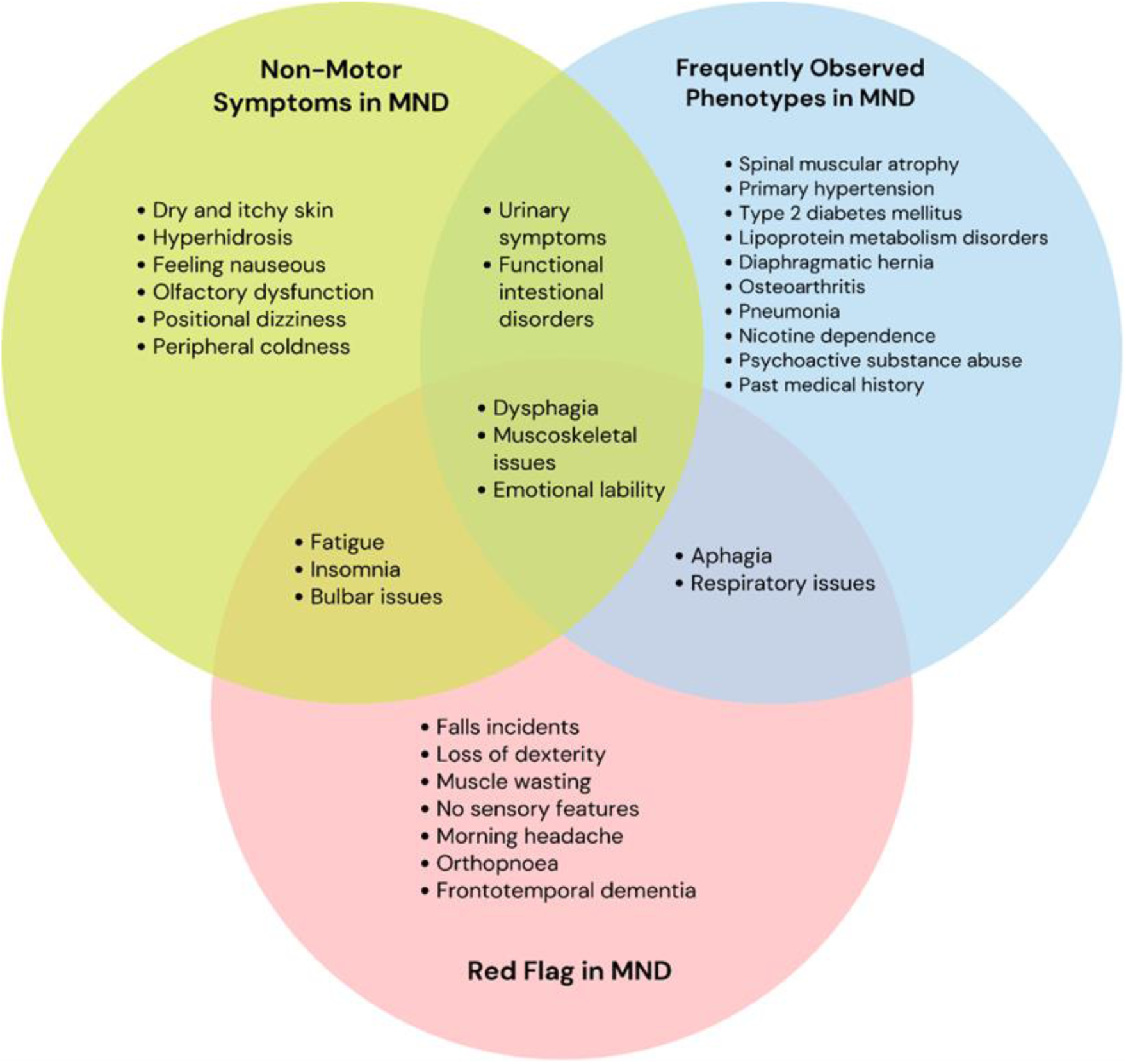
Venn diagram illustrating the overlap between non-motor symptoms in amyotrophic lateral sclerosis (ALS), frequently phenotypes observed in individuals with motor neurone disease (MND) before diagnosis, and the MND Red Flag List. Non-motor symptoms (blue) include sensory disturbances, dizziness, and autonomic dysfunction. Frequent phenotypes (pink) represent comorbid conditions and prior diagnoses identified in real-world data, including metabolic disorders and hypertension. The MND Red Flag List (green) highlights key clinical indicators for early diagnosis, such as muscle wasting, loss of dexterity, and frontotemporal dementia. Overlapping regions indicate shared symptoms across categories, including fatigue, bulbar dysfunction, and respiratory issues.

**Figure S10:**
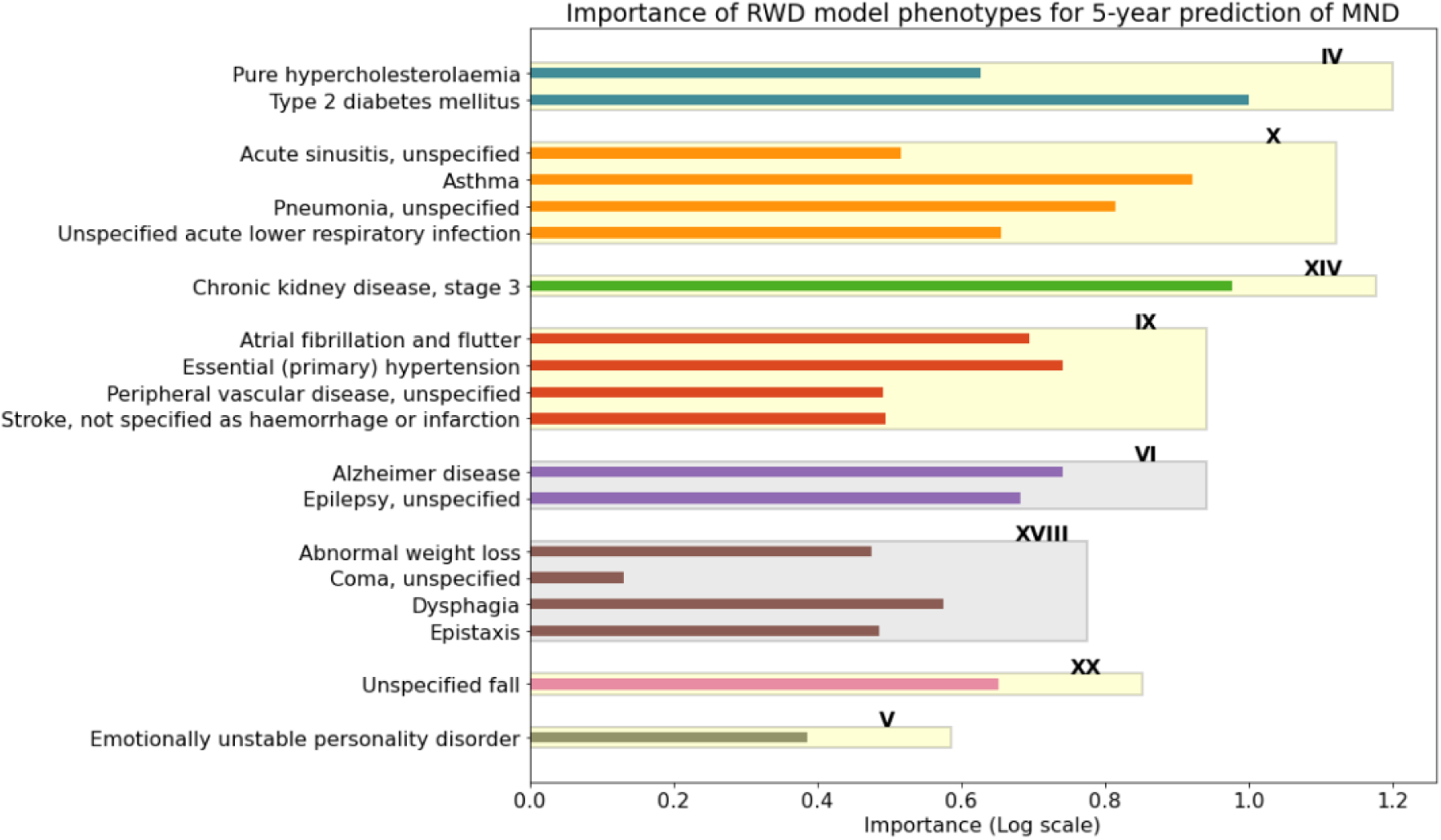
This figure presents the relative importance of features used in the predictive modelling of MND 5 years before diagnosis for the ensemble approach for the Red Flag List, Graph Features and Real-World Data. Feature importance was derived from real-world data (HES) and reflects the contribution of individual clinical variables— mapped to ICD-10 codes and grouped by chapters—to model performance. The distribution underscores which features most strongly influence predictions, offering insights into potential early indicators and key factors associated with MND diagnosis.

